# The closed eye harbors a unique microbiome in dry eye disease

**DOI:** 10.1101/2020.01.08.20016865

**Authors:** Kent A. Willis, Cameron K. Postnikoff, Amelia B. Freeman, Gabriel Rezonzew, Kelly K. Nichols, Amit Gaggar, Charitharth V. Lal

## Abstract

Dry eye affects millions of individuals. In experimental models, dry eye disease is associated with T helper cell 17-mediated inflammation of the ocular surface that may cause persistent damage to the corneal epithelium. However, the initiating and perpetuating factors associated with chronic inflammation of the ocular surface remain unclear. The ocular microbiota alters ocular surface inflammation and may influence dry eye disease development and progression. Here, we collected serial samples of closed eye tears during a randomized clinical trial of a non-pharmaceutical dry eye therapy and used 16S rRNA metabarcoding to characterize the microbiome. We show the closed dry eye microbiome is distinct from the healthy closed eye microbiome. The ocular microbiome was described only recently, and this report implicates a distinct microbiome in ocular disease development. Our findings suggest an interplay between microbial commensals and inflammation on the ocular surface. This information may inform future studies of the pathophysiological mechanisms of dry eye disease.

## Introduction

Dry eye disease is a common condition that affects millions of individuals worldwide^1,2^. Ocular surface inflammation has recently been recognized as a hallmark of dry eye disease^3,4^. T helper type 17 (Th17) cells on the inflamed ocular surface mediate the long-term progression of dry eye disease in experimental models^3^, however, this same pathophysiology has yet to be demonstrated conclusively in humans. In chronic dry eye, persistent inflammation may eventually produce lasting corneal epithelial damage^4^. However, the factors that incite and perpetuate inflammation in dry eye disease remain unknown^5^. Microbial commensal organisms can alter Th17 populations in the host organism, both in homeostasis and when perturbed^6,7^. In contrast to previous beliefs, the human eye hosts a resident microbiome^8-11^. Culture and molecular-based swabs of the ocular surface have revealed an intrinsic microbiome distinct from the surrounding skin, but information about the tear microbiome is scarce^12^. Interestingly, the ocular microbiome impacts ocular surface inflammation^13^. Together, these findings suggest the ocular microbiome may cause or perpetuate the development of chronic dry eye. Here, we collected longitudinal samples of the bilateral closed eye tear microbiome from a randomized clinical trial for the treatment of dry eye disease. We aimed to identify a distinct closed eye microbiome in dry eye disease. We hypothesized that the chronic dry eye microbiome, if present, would be distinct from the normal closed eye microbiome.

## Results

### Cohort

Based on the power and pairwise sample-size estimator for permutational multivariate analysis of variance (PERMANOVA) application Micropower^14^, a low abundance 16S rRNA dataset similar to previous studies of the ocular micribome^12^ would require a minimal sample size of 30 to generate a discriminatory power of 0.8 with a significance level of 0.05. Therefore, we aimed for at least 35 subjects per study arm. **Table** 1 shows the demographics and clinical characteristics of the enrolled participants. Based on a post-enrollment exam and two clinical surveys, after informed consent, participants were classified as dry eye or controls (**Supplemental S1**).

The resulting 36 control and 36 dry eye subjects were then stratified by clinical severity of eye disease (**Figure 1** and **Supplemental Table 1**). The study arms were randomized to daily saline eye wash upon awakening or non-intervention. A baseline closed eye tear sample was collected at randomization and the final sample after one month, yielding 144 samples. We observed no differences within the normal and dry eye cohorts (normal vs mild, or moderate vs severe). Therefore, all subsequent analyses were performed on normal or dry eye only (**Methods**).

**Table 1.** Participant demographics and clinical characteristics.

|  | Normal Eye | Dry Eye | P value |
| --- | --- | --- | --- |
| Subjects, n | 36 | 36 |  |
| Sex |  |  | 1.00 |
| Female, n (%) | 23 (64) | 23 (64) |  |
| Male, n (%) | 13 (36) | 13 (36) |  |
| Age, y ± SD | 33 ± 11 | 52 ± 19 | <0.0001 |
| DEQ Score | 1.2 ± 1.3 | 12 ± 3.3 | <0.0001 |
| Phenol Red Thread | 22 ± 5.3 | 19 ± 8.6 | 0.08 |
| Wetting length, mm ± SD | | | |
| NIK BUT, s ± SD | 15 ± 5.2 | 10.7 ± 5.2 | 0.0007 |
| Corneal Staining | 0.47 ± 0.8 | 3.6 ± 3 | <0.0001 |
| InflammaDry Positive, n (%) | 13 (36) | 20 (55) | 0.10 |
| Randomization |  |  |  |
| Control, n (%) | 16 (44) | 20 (56) |  |
| Treatment, n (%) | 20 (56) | 16 (44) |  |

**Fig. 1.**
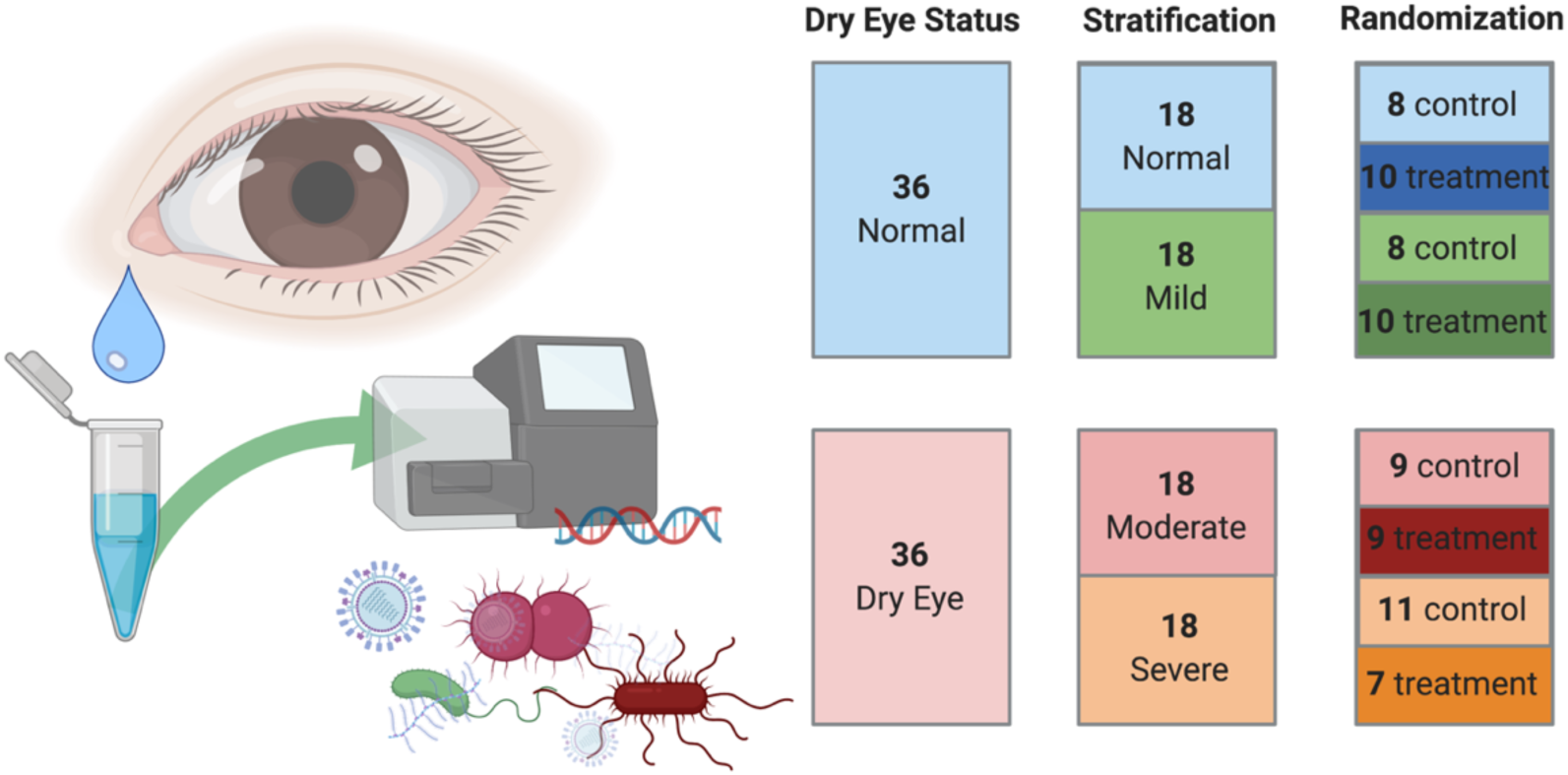
Subjects were allocated, stratified and randomized to treatment or control. Schematic of collection of closed eye tears in sterile saline, followed by 16S rRNA metabarcoding of the bacterial microbiome. Diagram of subject allocation and randomization. Figure generated with BioRender.

### Sequencing data

To investigate the bacterial microbiome of the closed dry eye, we used metabarcoding of the 16S rRNA gene. Sequence reads were generated using the Illumina MiSeq system (57,022 ± 47,011 counts/sample, **Methods**). We assigned taxonomy using the Greengenes database^15^, producing 1,593 data points/sample. After excluding any reads aligned to chloroplasts or cyanobacteria, we examined the bacterial community composition using the remaining 1,195 data points/sample that were collectively assigned to 185 genera^16^. All tear samples had the standard > 1000 aligned reads/sample, so we retained all samples for further analysis. We used a log_2_-transformation of cumulative sum scaling^17^ (log_2_ CSS) to normalize our dataset, consistently producing 8,700-10,000 reads per sample (**Supplemental Fig. S2**).

### The closed eye microbiome is distinct in dry eye disease

In the taxonomic analysis of the closed eye microbiome, the microbiomes of individuals with chronic dry eye disease and those without form distinct communities. Dry eye microbial communities are more diverse as quantified by richness (ANOVA, *f* =4.8, *P* =7.5 x10^−5^), evenness (ANOVA, *f* =13, *P* =2.1×10^−12^) and Shannon diversity (ANOVA, *f* = 12, *P* =5.9 x10^−12^, **Fig. 2a**). As demonstrated by principal coordinate analysis (PCoA) of Bray-Curtis dissimilarity, these communities form unique clusters as quantified by PERMANOVA (*R*^*2*^ =0.21, *P* =0.00033, **Fig. 2b**). Similarly, multivariate redundancy analysis (RDA, variance =95.25, *f* =3.71, *P* =0.001) and canonical correspondence analysis (CCA, chi^2^ =0.16 *f* =3.64, *P* =0.001) showed these communities are distinct, and this factor accounts for the majority of variance in the data. As quantified by two-way ANOVA adjusted for false discovery rate (FDR), univariate analysis of the relative abundance of bacterial genera showed individuals with dry eye disease have differences in the relative abundance of 113 genera, with the most significant differences in *OPB56, Methylobacteriaceae, Bacteroidetes, Pseudomonas*, and *Meiothermus* (all with FDR-adjusted *P* < 2 x10^−22^, **Supplemental Table 2**). Similarly, mixed effects regression detected the most significant differences in the abundance of *OPB56, Bacteroidetes, Pseudomonas, Meiothermus*, and *Methylobacteriaceae* (all with FDR-adjusted *P* < 3.1 x10^−20^, **Supplemental Table 3**) among 49 genera with significant differences in relative abundance. **Fig. 2c** shows relative abundance at the order level. To further investigate the importance of particular bacterial operational taxonomic units (OTUs), we used Spearman network analysis (**Fig. 2d**).

**Fig. 2.**
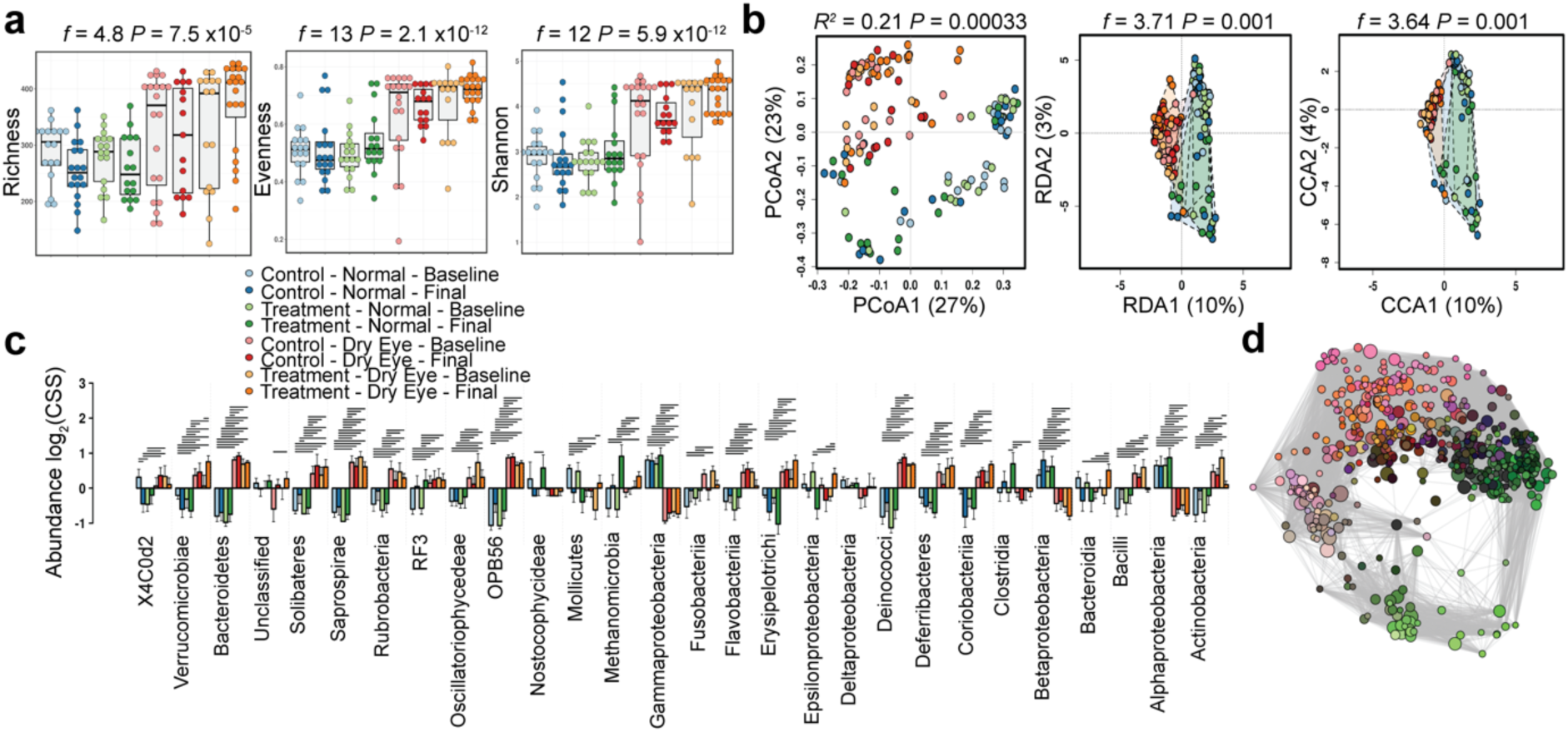
Patients with dry eye have a different closed eye microbiome. (a) Individuals with chronic dry eye have different alpha diversity of the tear microbiome as quantified by the richness (ANOVA, *f* =4.8, *P* =7.5 ×10^−5^), evenness (ANOVA, *f* =13, *P* =2.1×10^−12^) and Shannon diversity (ANOVA, *f* = 12, *P* =5.9 x10^−12^) indices. (b) The beta diversity of the closed eye microbiome in individuals with dry eye disease is distinct by principal coordinate analysis (PCoA) of Bray-Curtis dissimilarity (*R*^*2*^ =0.21 *P* =0.00033), redundancy analysis (RDA, variance =95.25, *f* =3.71, *P* =0.001) and canonical correspondence analysis (CCA, chi^2^ =0.16 *f* =3.64 *P* =0.001). (c) Relative abundance of bacterial orders. Log_2_(CSS), Log_2_ transformation of cumulative-sum scaling. Two-way ANOVA, * *P* =0.01, ** *P* =0.001, *** *P* = 0.0001. (d) Spearman network analysis at the OTU level. Positive correlations with a *P*-value < 0.05 are shown as an edge with the relative size determined by the importance of the taxa to the network.

### The closed dry eye microbiome remains distinct despite daily eye wash

Daily eye wash on awakening with sterile saline is a proposed therapy for dry eye disease^18^. We tested if the microbial community composition could be normalized by the prescription of daily saline eye wash. However, the microbial communities of dry (**Supplemental Fig. S3**) and normal eyes (**Supplemental Fig. S4**) were relatively unaffected by daily eye wash. The diversity of subjects with dry eye remained higher than that of normal individuals (richness, ANOVA, *f* =9.2, *P* =7.5 x10^−5^, evenness, ANOVA, *f* =28, *P* =2.8 x10^−14^ and Shannon diversity, ANOVA, *f* =28, *P* =6.1 x10^−14^, **Fig. 3a**). Multivariate analysis of these communities also remained distinct (PCoA, PERMANOVA, *R*^*2*^ =0.15, *P* =0.00033, RDA, variance =64.75, *f* =5.73, *P* =0.001 and CCA, chi^2^ =0.11, *f* =5.62, *P* =0.001, **Fig. 3b**). Univariate analysis detected the most significant differences in the abundance of *MLE112, Lactobacillaceae, Streptococcus, Sphingobium, Caldicoprobacter* and *Anaerococcus* (ANOVA, *P* < 0.01, **Fig. 3c**). Core microbiome analysis highlighted key differences in the distribution of key genera, and network analysis demonstrated the importance of key taxa (**Fig. 3d, 3e**).

**Fig 3.**
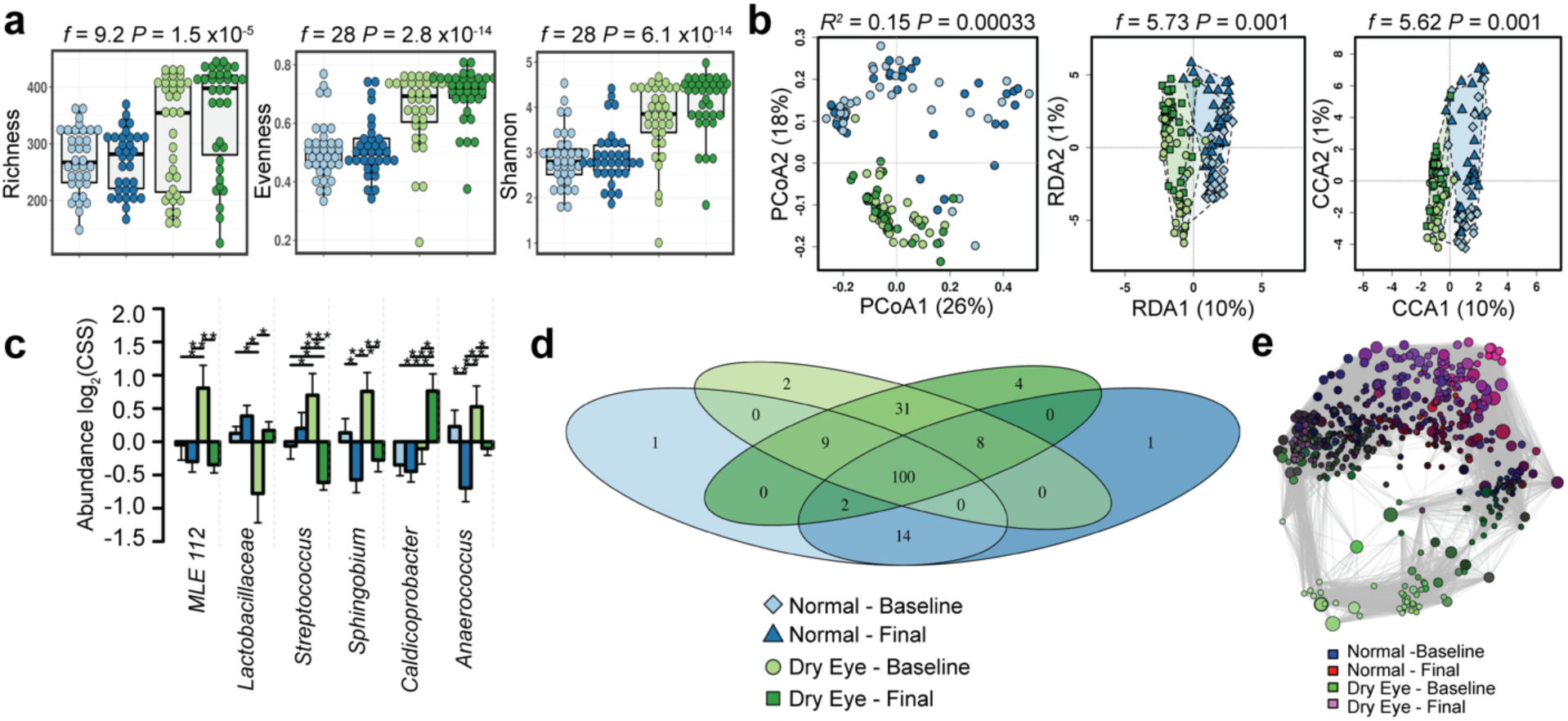
Daily eye rinse does not alter the closed eye microbiome. (a) Despite daily eye washes, the alpha diversity of the closed eye microbiome remains similar to baseline in individuals with and without dry eye disease as quantified by the richness (ANOVA, *f* =9.2, *P* =7.5 x10^−5^), evenness (ANOVA, *f* =28, *P* =2.8 x10^−14^) and Shannon diversity (ANOVA, *f* =28, *P* =6.1 x10^−14^) indices. (b) The beta diversity of the closed eye microbiome remains distinct in individuals with and without dry eye disease by principal coordinate analysis (PCoA) of Bray-Curtis dissimilarity (PERMANOVA, *R*^*2*^ =0.15 *P* =0.00033), redundancy analysis (RDA, variance =64.75, *f* =5.73, *P* =0.001) and canonical correspondence analysis (CCA, chi^2^ =0.11 *f* =5.62 *P* =0.001). (c) Relative abundance of bacterial genera. Log_2_(CSS), Log_2_ transformation of cumulative-sum scaling. Two-way ANOVA, * *P* =0.01, ** *P* =0.001, *** *P* = 0.0001. (d) Core microbiome analysis showing differences in microbial colonization at the genus level. (e) Spearman network analysis at the OTU level. Positive correlations with a *P*-value < 0.05 are shown as an edge with the relative size determined by the importance of the taxa to the network.

### The closed eye microbiome in dry eye disease is distinct at baseline

To gain further insight into the microbial community composition of the dry eye, we narrowed our focus to examine the closed eye microbial composition at the time of randomization. As indicated by our more general analysis, both diversity (richness, ANOVA, *f* =6.1, *P* =0.016, evenness, ANOVA, *f* =24, *P* =5.8 x10^−6^ and Shannon diversity, ANOVA, *f* =23, *P* =8.1 x10^−8^, **Fig. 4a**) and multivariate clustering of these communities are distinct at baseline (PCoA, PERMANOVA, *R*^*2*^ =1.34 *P* =0.00033, RDA, variance =53.95, *f* =7.12, *P* =0.001 and CCA, chi^2^ =0.09, *f* =7.12, *P* =0.001, **Fig. 4b**). Univariate analysis using negative binomial regression identified *Methylobacteriaceae, Pseudomonas, Bradyrhizobium* and *Allobaculum* as the five most differently abundant genera (all with FDR-adjusted *P* < 1.1 x10^−11^, **Supplemental Fig. S5a** and **Supplemental Table 4**). At the phylum level, Firmicutes and Bacteroidetes remained unchanged, while Verrucomicrobia (FDR *P* =4.6 x10^−11^) and Proteobacteria (FDR *P* =6.0 x10^−10^) showed the most significant differences. Core microbiome analysis identified 45 genera unique to dry eye with only 14 genera unique to the normal eye (**Fig. 4d**). Discriminant analysis of principal components at the order level revealed the abundances of orders OPB56 and Rhizobiales are important to discriminate the normal eye while the orders Halanaerobiales, Erysipelotrichales and Anaeroplasmatales are important to discriminate dry eye (**Fig. 4e**). Network analysis provided further evidence of these distinct communities (**Fig. 4f**).

**Fig. 4.**
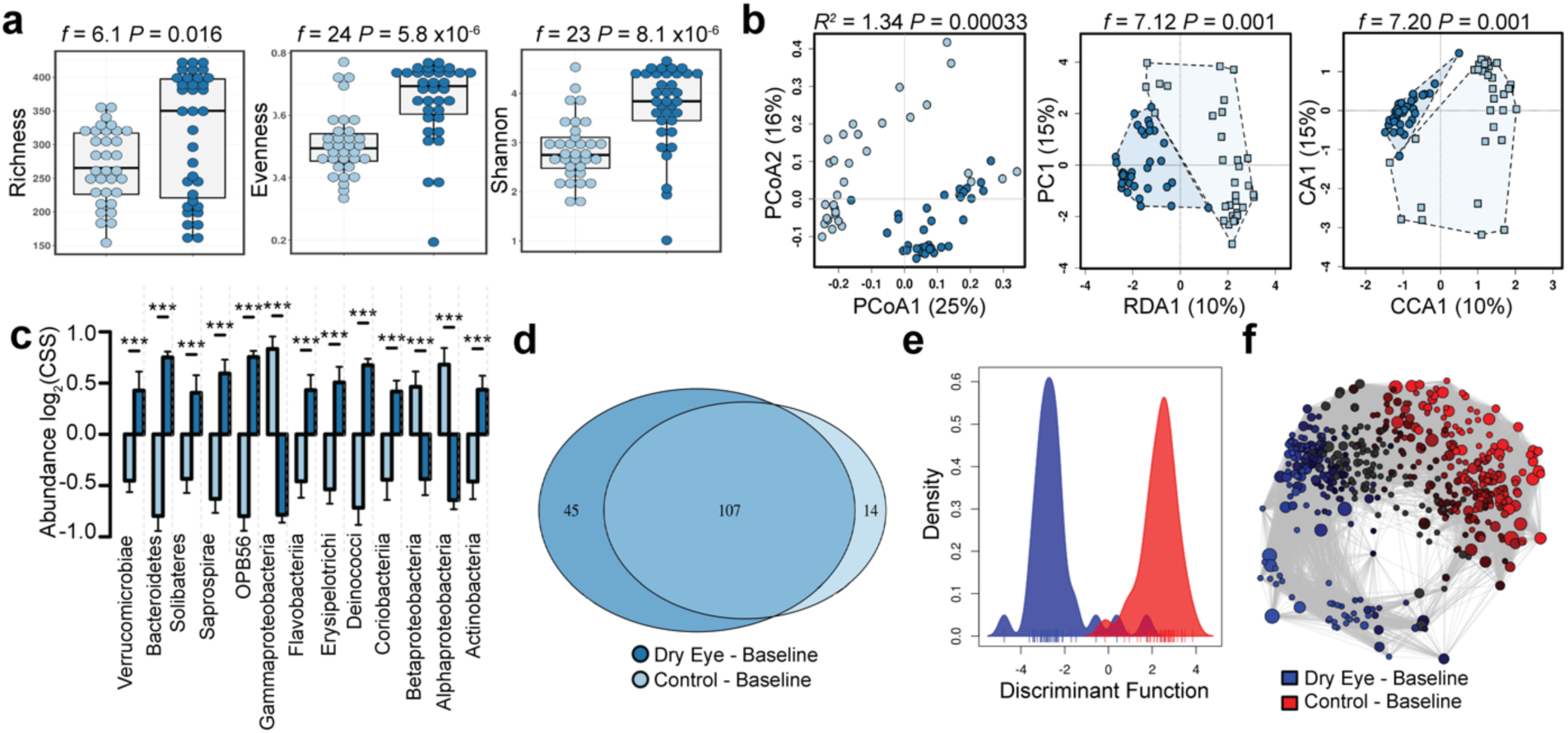
The closed eye microbiome of the in dry eye disease is distinct at baseline. (a) The alpha diversity of the closed eye microbiome remains distinct as quantified by the richness (ANOVA, *f* =6.1, *P* =0.016), evenness (ANOVA, *f* =24, *P* =5.8 x10^−6^) and Shannon diversity (ANOVA, *f* =23, *P* =8.1 x10^−8^) indices. (b) The beta diversity of the closed eye microbiome remains distinct as quantified by principal coordinate analysis (PCoA) of Bray-Curtis dissimilarity (PERMANOVA, *R*^*2*^ =1.34 *P* =0.00033), redundancy analysis (RDA, variance =53.95, *f* =7.12, *P* =0.001) and canonical correspondence analysis (CCA, chi^2^ =0.09, *f* =7.12, *P* =0.001). (c) Relative abundance of bacterial orders. Log_2_(CSS), Log_2_ transformation of cumulative-sum scaling. Two-way ANOVA (***displaying only, *P* < 0.001). (d) Core microbiome analysis showing differences in microbial colonization at the genus level. (e) Discriminant analysis of principal components at the order level. (f) Spearman network analysis at the OTU level. Positive correlations with a *P*-value < 0.05 are shown as an edge with the relative size determined by the importance of the taxa to the network.

### Machine learning accurately classifies dry eye samples at baseline

To more precisely identify unique features with the potential to function as biomarkers for patients with dry eye, we used linear discriminant analysis of effect size. We identified 5 genera that reliably identified individuals with dry eye (*Pseudomonas, Methylobacteriaceae, Helicobacter, Acetobacter* and *Stenotrophomonas*) with 3 genera that reliably identified normal patients (*Leuconostocaceae, Streptococcus* and *Calothrix*, **Supplemental Fig. S5b**). To further support the uniqueness of microbial communities in dry eye, we built a support vector machine using leave-one-out cross-validation that could identify samples with 94% accuracy. Similarly, when we developed a random forest classifier, variable importance analysis identified the prevalence of the genera *Methylobacterium, Megasphaera, Parabacteroides, S247, Bifidobacterium, Streptococcus, Desulfovibrio, Acetobacter, Dialister* and *Bacillus* as particularly useful to identify samples from individuals with dry eye (Importance > 20, **Supplemental Fig. S5c**).

### The closed eye microbiome in dry eye disease diverges after one month

We examined the stability of the dry eye microbiome by focusing on samples collected one month later in the same individuals. The dry eye microbiome remained distinct from the normal microbiome and remained consistent with the community composition noted at baseline (**Supplemental Fig. S3, S2**). Only slight divergence was noted over the course of a month (**Fig. 5**). The diversity of dry eye remained higher than that of the normal eye (richness, ANOVA, *f* =19, *P* =5.2 x10^−5^, evenness, ANOVA, *f* =65, *P* =1.9 x10^−11^ and Shannon diversity, ANOVA, *f* =60, *P* =8.6 x10^−11^, **Fig. 5a**). The distinct clustering of these communities on multivariate analysis also persisted (PCoA of Bray-Curtis dissimilarity, PERMANOVA, *R*^*2*^=1.52, *P* =0.00033, RDA, variance =66.53, *f* =8.52, *P* =0.001 and CCA, chi^2^ =0.10, *f* =8.14, *P* =0.001, **Fig. 5b**). Similarly, negative binomial regression noted the greatest differences in the abundance of the genera *Methylobacteriaceae, Pseudomonas, Azospirillum, Bradyrhizobium* and *Coriobacteriaceae* (all with FDR-adjusted *P* < 7.0 x10^−18^, **Supplemental Fig. S6a, Supplemental Table 4**). However, core microbiome analysis detected 76 unique genera in dry eye, 69 shared genera and 24 genera unique to the normal eye (**Fig 5d**). Similar to baseline, discriminant analysis of principal components at the order level identified OPB58 and Bacteroidetes as important discriminators of the normal eye. In individuals with dry eye, additional orders enabled further discrimination, with Flavobacteriales, Alteromonadales and Actinomycetes, Anaeroplasmatales and Desulfuromonadales functioning as the primary discriminators instead of Halanaeobiales and Erysipelotrichales (**Fig. 5d**).

**Fig 5.**
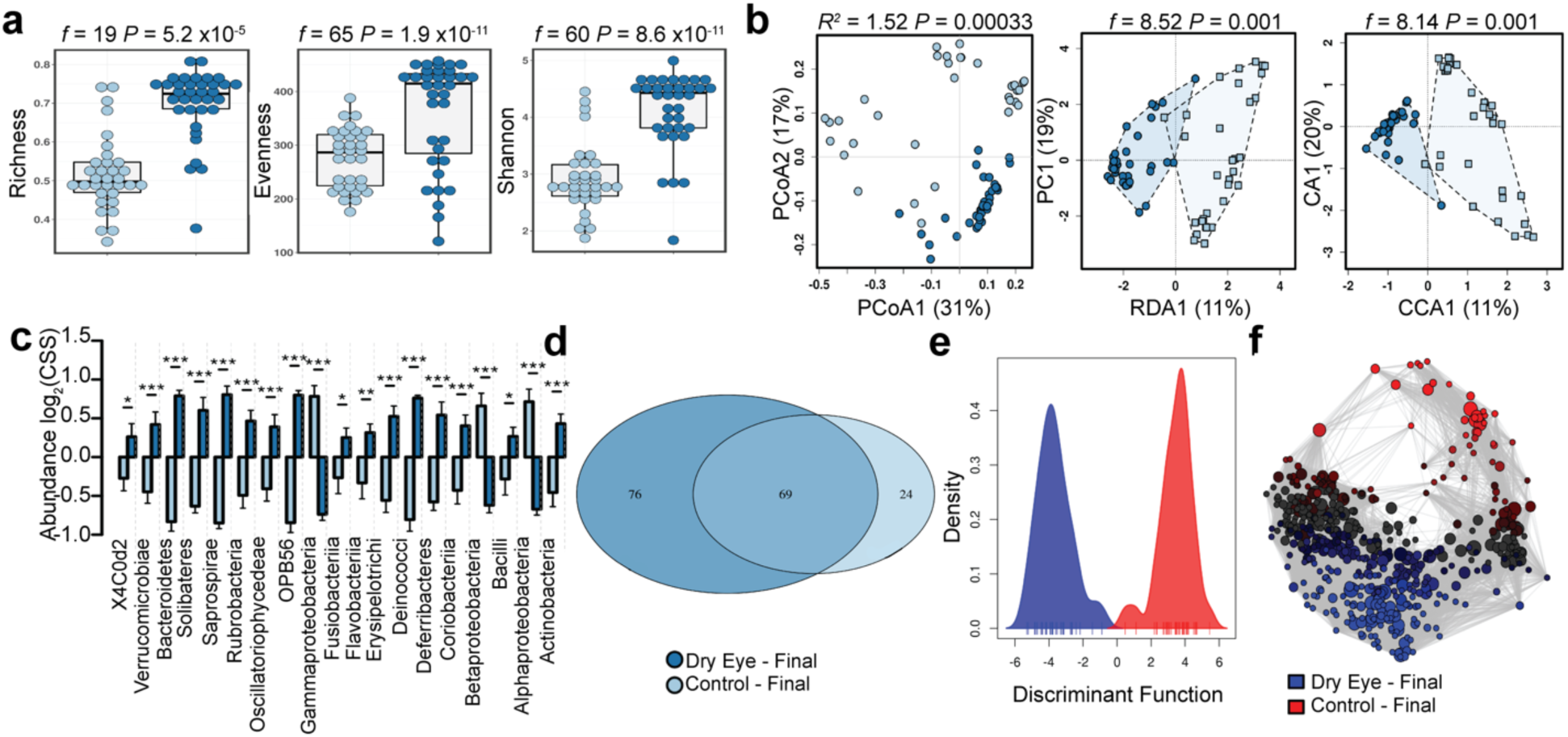
The closed eye microbiome in dry eye disease remains distinct. (a) The alpha diversity of the closed eye microbiome remains distinct as quantified by the richness (ANOVA, *f* =19, *P* =5.2 x10^−5^), evenness (ANOVA, *f* =65, *P* =1.9 x10^−11^) and Shannon diversity (ANOVA, *f* =60, *P* =8.6 x10^−11^) indices. (b) The beta diversity of the closed eye microbiome remains distinct as quantified by principal coordinate analysis (PCoA) of Bray-Curtis dissimilarity (PERMANOVA, *R*^*2*^ =1.52 *P* =0.00033), redundancy analysis (RDA, variance =66.53, *f* =8.52, *P* =0.001) and canonical correspondence analysis (CCA, chi^2^ =0.10, *f* =8.14, *P* =0.001). (c) Relative abundance of bacterial orders. Log_2_(CSS), Log_2_ transformation of cumulative-sum scaling. Two-way ANOVA, * *P* =0.05, ** *P* =0.01, *** *P* = 0.001. (d) Core microbiome analysis showing differences in microbial colonization at the order level. (e) Discriminant analysis of principal components at the order level. (f) Spearman network analysis at the OTU level. Positive correlations with a *P*-value < 0.05 are shown as an edge with the relative size determined by the importance of the taxa to the network.

### Machine learning more accurately identifies dry eye after a month

We used linear discriminant analysis of effect size to identify bacterial genera with the potential function as biomarkers and noted 10 genera unique to individuals with dry eye and 9 unique to individuals with a normal eye, supporting the limited divergence of these communities (**Supplemental Fig. S6b**). We also developed a support vector machine with leave-one-out cross-validation that identified dry eye samples with 97% accuracy. Variable importance analysis of a random forest classifier noted the prevalence of *Methylobacterium, Megasphaera, Parabacteroides, S247, Bifidobacterium, Streptococcus, Desulfovibrio* and *Acetobacter* as important identifiers (Importance > 20, **Supplemental Fig. S6c**).

### The closed eye microbiome is unaltered by the cellular concentration of the tear fraction

To ensure our microbial ecological analysis was unaltered by the cellular content of the tear fraction, we compared the microbial composition of each cellular fraction collected. Three samples were excluded from this analysis due to incomplete data (n = 141). Additionally, community composition was similar when we collated high and low cellular fractions to increase discriminatory power (**Fig. 6**).

**Fig. 6.**
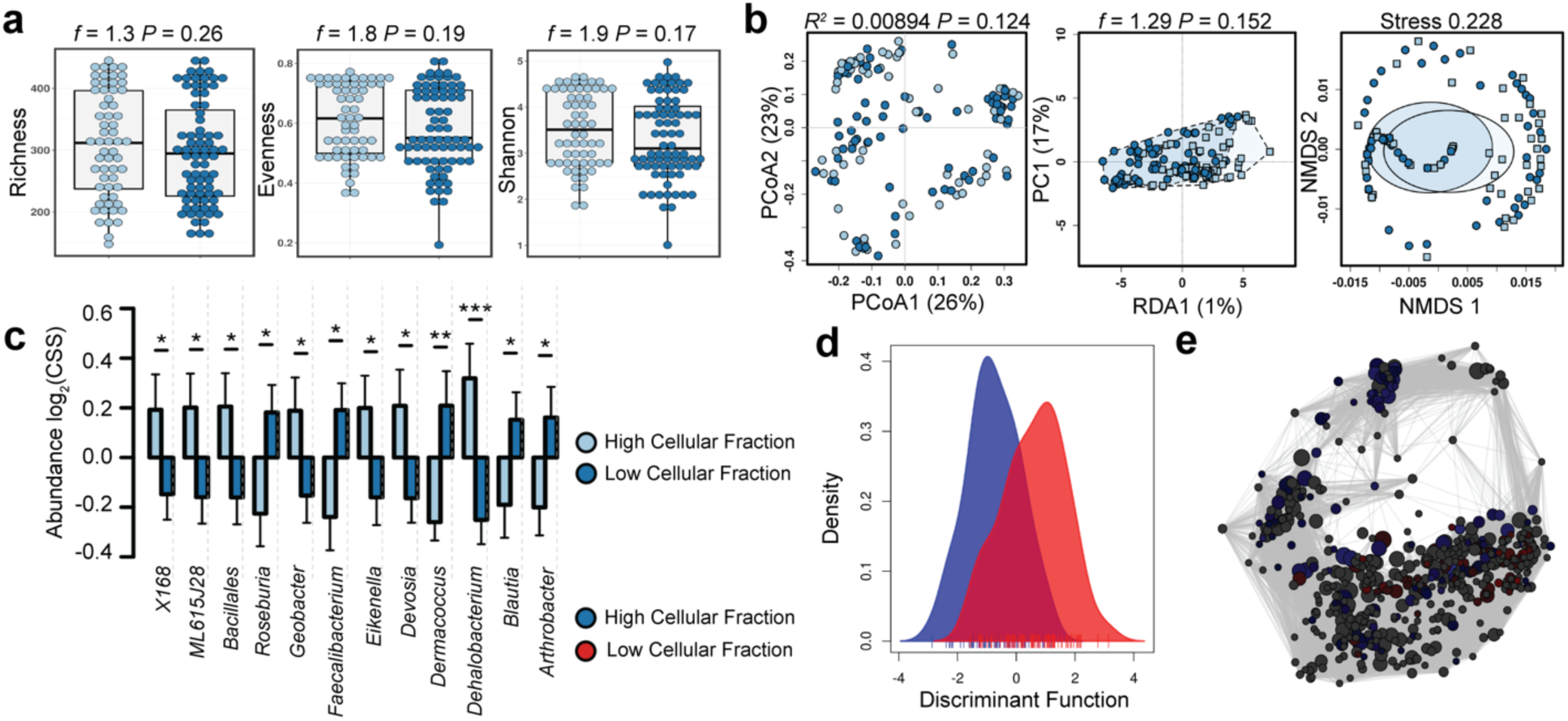
The closed eye microbiome is unaltered by the cellular fraction. a) The alpha diversity was not different as quantified by the richness (ANOVA, *f* =1.3, *P* =0.26), evenness (ANOVA, *f* =1.8, *P* =0.19) or Shannon diversity (ANOVA, *f* =1.9, *P* =0.17) indices. (b) The beta diversity of the closed eye microbiome is unaltered by the cellular fraction as quantified by principal coordinate analysis (PCoA) of Bray-Curtis dissimilarity (PERMANOVA, *R*^*2*^ =0.00894 *P* =0.124), redundancy analysis (RDA, variance =4.82, *f* =1.29, *P* =0.152) and non-metric multidimensional scaling (NMDS, stress = 0.228). (c) Relative abundance of bacterial genera. Log_2_(CSS), Log_2_-transformation of cumulative-sum scaling. Two-way ANOVA, * *P* =0.05, ** *P* =0.01, *** *P* =0.001. (d) Discriminant analysis of principal components. (e) Spearman network analysis at the OTU level. Positive correlations with a *P*-value < 0.05 are shown as an edge with the relative size determined by the importance of the taxa to the network.

## Discussion

Direct crosstalk between resident microbes and host immune cells at the mucosal surface is a critical determinant of inflammatory diseases^19^. Furthermore, Th17 and Treg cells are implicated in the development of chronic dry eye disease^3^ and particularly attuned to the ecology of the resident microbiota^6^. Therefore, the ocular surface microbiome may contribute to the pathogenesis of dry eye disease^13^. Understanding how the ocular microbiome influences ocular surface disease will be important for therapeutic development. In this report, we present initial evidence of a distinct closed eye microbiome in dry eye. Moreover, we provide the first evidence linking microbiome changes with ocular surface disease.

In this study, human subjects collected samples of sterile saline rinse applied to the ocular surface upon awakening in the morning. Using saline eye wash to collect microbiome samples is novel and less invasive than swabs of the ocular surface, which has been routinely used for the collection of tear leukocytes^18,20^. In a series of pooled samples from both eyes, the closed eye tear microbiome in individuals with dry eye disease was distinct from that in healthy controls. Compared to swabs, where the fiber content and limited sample area artificially reduce the number of detectable microbes^21^, saline wash may allow for a less biased sampling of the entire ocular surface, especially when pooled from both eyes. Furthermore, a general weakness of previous ocular microbiome studies is focusing on shifts in percent relative abundance to identify differences in bacterial taxa. A strength of our analysis is the rigorous log_2_ normalization of the cumulative sum scaling transformation performed prior to analysis. With this normalization, our data more accurately reflect true shifts in relative abundance^17^. Using untransformed data, the most significant difference in our dataset was a nearly complete substitution of *Staphylococcus* spp. for *Pseudomonas* spp. in patients with dry eye. Doan *et al*. reported a similar shift in the unnormalized relative abundance of *Staphylococcus* spp. and *Pseudomonas*^22^. A more rigorous normalization performed on our dataset confirmed the reduction in *Pseudomonas* spp. but not in *Staphylococcus* spp. In addition, 48 other genera may warrant further investigation.

Previous research has focused on the open eye microbiome^12^, while we focused on the closed eye microbiome. Every night during sleep, inflammatory species move into the closed eye tear film^23,24^, perhaps in response to entrapped microbiota^12,25^. If microbial abundance increases in this situation, differences in bacterial community composition could become more pronounced in the closed eye microbiome than in the open eye. This difference could lead to the closed eye producing more reliable markers of disease states for diagnostic development. Alternatively, this difference could help determine whether anti-inflammatory mechanisms in the closed eye are sufficient to maintain a healthy ocular microbiome.

Graham and colleagues explored the microbiome of dry eye using culture and broad 16S rRNA-based PCR^23^. Due to considerable differences in next generation sequencing and bioinformatics techniques in the last decade as well as our different sampling methods, directly comparing our 16S rRNA-based characterizations of the ocular microbiome is difficult. Nevertheless, our analysis does support their findings of increased bacterial diversity in patients with dry eye disease^23^. Although we identified a broader number of genera than was feasible when Graham and colleagues^23^ performed their analysis, we confirmed their identification of several genera in dry eye, including *Streptococcus, Staphylococcus, Corynebacterium* and *Propionibacterium*. They nevertheless detected more colony forming units in patients with dry eye than in controls using culture-based techniques. These techniques are biased in low biomass samples because of the limited growth of some species in culture^12^. Coagulase negative *Staphylococcus* spp. were the most commonly identified taxa via culture, and their PCR-based analysis showed the microbiome remained stable over three months. Another culture-based pilot study identified similar increased abundances of *Staphylococcus aureus*, coagulase negative *Staphylococcus, Corynebacterium* and *Propionibacterium* in chronic dry eye^24^. We also provided in-depth analysis of population dynamics and demonstrated possible divergence of microbial diversity and community structure of normal and dry eyes over time.

Increased bacterial diversity is considered favorable, as more diverse communities are often more resistant to perturbation^25^. However, mounting evidence suggests the ocular microbiome deviates from this trend, likely due to lysozyme and antimicrobial compounds in tears^26^. In dry eye disease, our results suggest increased microbial diversity is a hallmark of disease. Consistent with the behavior of the ocular microbiome in other pathological conditions^12^, this finding may represent breakdown of the microbe-oriented homeostatic mechanisms of the host. This finding may also suggest increased diversity results from the incorporation of pathobionts into this community. Alternatively, the altered ecology may result from changes within the ocular surface of the dry eye that support establishment of a wider diversity of microbes in this environment. These additional species may not be harmful but instead highlight the dynamic biome within the human eye. The evidence may also suggest the immune system of the closed eye cannot regulate the ocular surface microbiome during sleep. In the closed eye, preliminary evidence suggests higher numbers of neutrophils with an enhanced degranulation response in dry eye disease^18,Postnikoff:2018fv; 20^, indicating a dysregulated immune response. This dysregulation may permit increased microbial diversity. The reverse could also be true: enhanced microbial diversity may induce a greater inflammatory response.

This dataset consistently demonstrates the microbial communities in dry eye disease are distinct and remain distinct even with daily saline eye wash. We asked subjects in the treatment arm to self-administer saline eye wash immediately upon awakening. We hypothesized this time may represent the point of maximum difference since microbes likely accumulate on the ocular surface overnight. However, we did not observe reduced bacterial diversity with saline eyewash, with dry eye communities becoming more similar to control eye communities with treatment. Instead, the communities slightly diverged: the microbial communities in dry eye remained different from those in healthy eyes. Thus, the mechanisms of microbial accumulation on the ocular surface may be more complex than moisture content or tear clearance. Intrinsic immune or mechanical alterations in dry eye disease may foster differences in commensal abundance, potentially explaining how altered bacterial communities on the ocular surface contribute to dry eye disease.

## Methods

### Participants

This study was performed in accordance to the Declaration of Helsinki under the supervision of the Institutional Review Board of the University of Alabama at Birmingham (UAB), Birmingham, AL, USA. We performed a secondary analysis of the closed eye microbiome from a prospective, randomized controlled trial of the efficacy of daily eye wash to ameliorate inflammation in individuals with dry eye disease (ClinicalTrials.gov NCT03332342). Future studies may more accurately localize the innate microbes of the eye by examining ocular swabs and the tear microbiome simultaneously. Serial sampling throughout a day may demonstrate diurnal variation in these bacterial communities. However, aside from certain antibacterial medications^20^, most human microbiome studies are observational by necessity. Gnotobiotic animals are often useful in unraveling causality in host-microbe interactions and could be the next step^27,28^, especially to explore interactions between the ocular microbiome and host Th17 and Treg cells in dry eye disease. Building on these experiments may include identifying changes in microbial metabolites or exploring how the gut microbial communities influence ocular development and disease. Finally, aside from one study that examined the virome^29-31^, most studies of the ocular microbiome have focused exclusively on bacteria. The fungal microbiome of the normal eye was described only recently^22^ and may warrant further investigation in dry eye disease. As performed recently for the gut^32^ and lung^33^, modeling the initial succession of multi-kingdom microbial communities in newborns may identify ecological factors in the ocular microbiome.

The ocular microbiome potentially hosts a distinct core microbiome that may be perturbed in eye disease^34^. Our study provides novel insight into the closed eye microbiome in dry eye disease. Th17 cells are sensitive to changes in the microbiome^6,7^ in experimental dry eye disease^3^. Therefore, increased microbial abundance in dry eye may promote the development of dry eye disease by altering T cell subpopulations on the ocular surface. More research is needed to reveal whether an altered microbiome drives the development of dry eye disease.

Informed consent was obtained from all participants prior to enrollment. Participants were selected based on the results of a physical exam and two clinical surveys, the Dry Eye Questionnaire 5 (DEQ5)^35^ and the Ocular Surface Disease Index (OSDI)^36^. Exclusion criteria included pregnancy, contact lens use within the past three months, glaucoma treatment, current tobacco use, systemic dry-eye-associated inflammatory disease, any anti-inflammatory therapy for dry eye treatment in the last three months or other antimicrobial or anti-inflammatory medications in the last month (**Supplemental Fig. S1**). Stratification into normal and dry eye was driven primarily by symptoms, as measured by the DEQ5 score, with further stratification among both groups determined by phenol red thread wetting length^37^, non-invasive tear break-up time (NIKBUT) and the InflammaDry test (Quidel; San Diego, CA, USA. **Supplemental Table 1**). Dry eye subjects also had to endorse the sensation of dry eye or have a previous diagnosis of dry eye disease. Equal-weight randomization to either daily eye wash or control was performed using REDCap (Research Electronic Data Capture; Nashville, TN, USA)^38^.

### Eyewash treatment

Subjects were trained on the first visit to perform a daily eye wash. Briefly, as described previously^39-41^, subjects were provided one 50 mL sterile centrifuge tube, and two 10 mL sterile polypropylene syringes (BD Biosciences; San Jose, CA, USA) with 5 mL of sterile saline per syringe. Immediately after awakening, subjects were instructed to wash their eyes by dispensing the saline gently across the ocular surface, with the runoff collected in a sterile Eppendorf tube. Runoff from both eyes was collected to produce a pooled sample of both eyes collected into the same tube. Subjects in the treatment group completed daily therapy for approximately 28 days.

### Sample collection

We utilized the power and pairwise sample-size estimator for PERMANOVA application Micropower^14^, as implemented at https://fedematt.shinyapps.io/shinyMB/, to estimate the minimum number of samples to produce a power of 0.8 to detect a difference with a significance level of 0.05 via PERMANOVA. We processed the collected eyewash samples immediately after they were dropped off by study subjects. Pooled tear collections were centrifuged at 270x g, and the supernatant was aliquoted into 1.5 mL microcentrifuge tubes and stored at −80°C. If more than one million cells were recovered in the pooled tear collection, supernatant aliquots were spiked with cells from the precipitate to increase the potential yield of microbiome analysis by normalizing the cellular fraction.

### Illumina MiSeq metabarcoding

In the Microbiome Core at UAB, microbial genomic DNA was isolated and PCR was used with unique bar-coded primers to amplify the V4 region of the 16S rRNA gene to create a 16S amplicon library of the samples. The PCR products were sequenced using the NextGen Illumina MiSeq System (Illumina Inc.; San Diego, CA, USA). The sequence data covered the 16S rRNA V4 region with a PCR product length of ∼255 bases and 250 base paired-end reads. The raw dataset is available at the NCBI Sequence Read Archive: http://www.ncbi.nlm.nih.gov/bioproject/597168.

### Bioinformatics

Sequences were grouped into OTUs using QIIME v1.9^42^. We assigned taxonomy using the Ribosomal Database Project classifier (threshold 0.8) against the Greengenes database^14,15^. We then imported these processed sequence reads into Calypso v8.84^43^ for further processing and data analysis. After excluding any OTUs assigned to chloroplasts or cyanobacteria, we *a priori* determined to exclude any samples with < 1000 reads/sample from downstream analysis. We then utilized cumulative sum scaling^17^ to normalize the relative abundance and performed a log_2_-transformation of the resulting data, centering the taxonomic counts to 0 and scaling to a range of - 2 to 2 with a variance of 1. For alpha diversity analysis, samples were rarified to a depth of 7804 reads/sample. Alpha diversity was then quantified using the richness, evenness and Shannon diversity indices^44^, testing for significant differences with ANOVA adjusted for FDR. For beta diversity, we visualized the data using PCoA, RDA, CCA or NMDS^45^. We used PERMANOVA to determine significant differences in beta diversity^46^. Relative abundance was quantified using ANOVA adjusted for FDR for multiple groups or negative binomial regression (DESeq2 function) for binary comparisons. For Spearman network analysis, we displayed positive correlations with an FDR-adjusted *P* < 0.05 as an edge. Relative size of the point represents the importance of the taxa to the resulting network. In addition, we used core microbiome analysis (of the 300 most abundant taxa), linear discriminant analysis of effect size, and discriminant analysis of principal components to select significant features. For analyses with two groups, we developed machine learning classifiers using support vector machine and random forest algorithms.

## Data Availability

The raw dataset is available at the NCBI Sequence Read Archive.

http://www.ncbi.nlm.nih.gov/bioproject/597168.

## Acknowledgements

The authors thank the Microbiome Resource at UAB for the performance of Illumina MiSeq. The following are acknowledged for their support of the Microbiome Resource: UAB School of Medicine, Comprehensive Cancer Center (P30AR050948), Center for AIDS Research (5P30AI027767), Center for Clinical Translational Science (UL1TR000165) and the Heflin Center. We also thank the Children’s Foundation Research Institute at Le Bonheur Children’s Hospital for providing scientific editing.

## Author Contributions

CKP, AG and CVL conceptualized the study. CKP, KN facilitated the performance of the randomized clinical trial and oversaw the sample collection. GR performed the DNA extraction. KAW designed and performed the biostatistical analyses, drafted the manuscript and prepared the figures. CVL directed the performance of the study and provided funding. KAW, CKP, AF, GR, KN, AG and CVL edited the manuscript and approved the final version.

## Additional Information

Supplementary information accompanies this preprint.

## Supplement

**Fig. S1.**
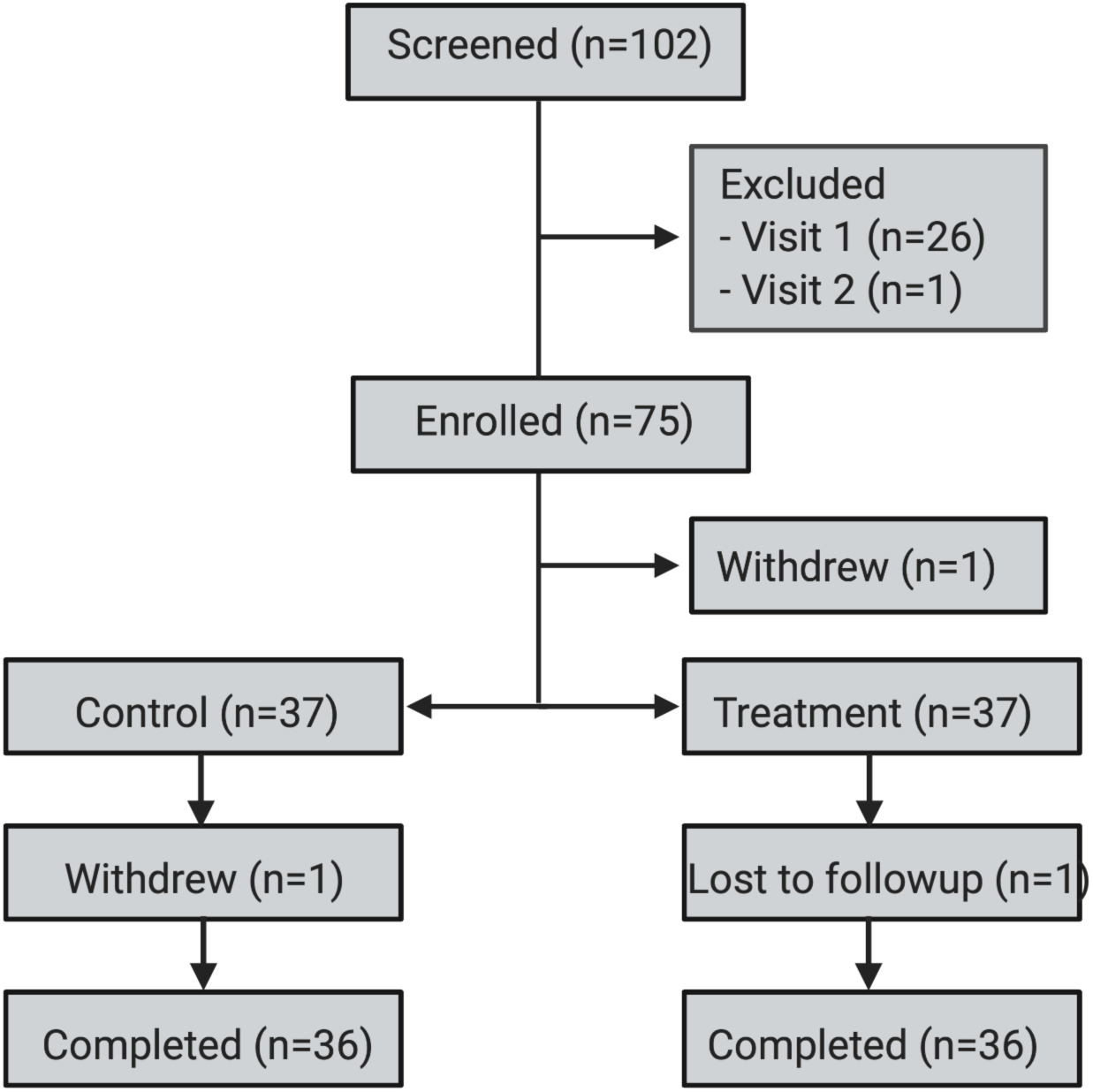
Consolidated standards of reporting trials subject allocation diagram. CONSORT subject allocation diagram. Figure generated by BioRender.

**Fig. S2.**
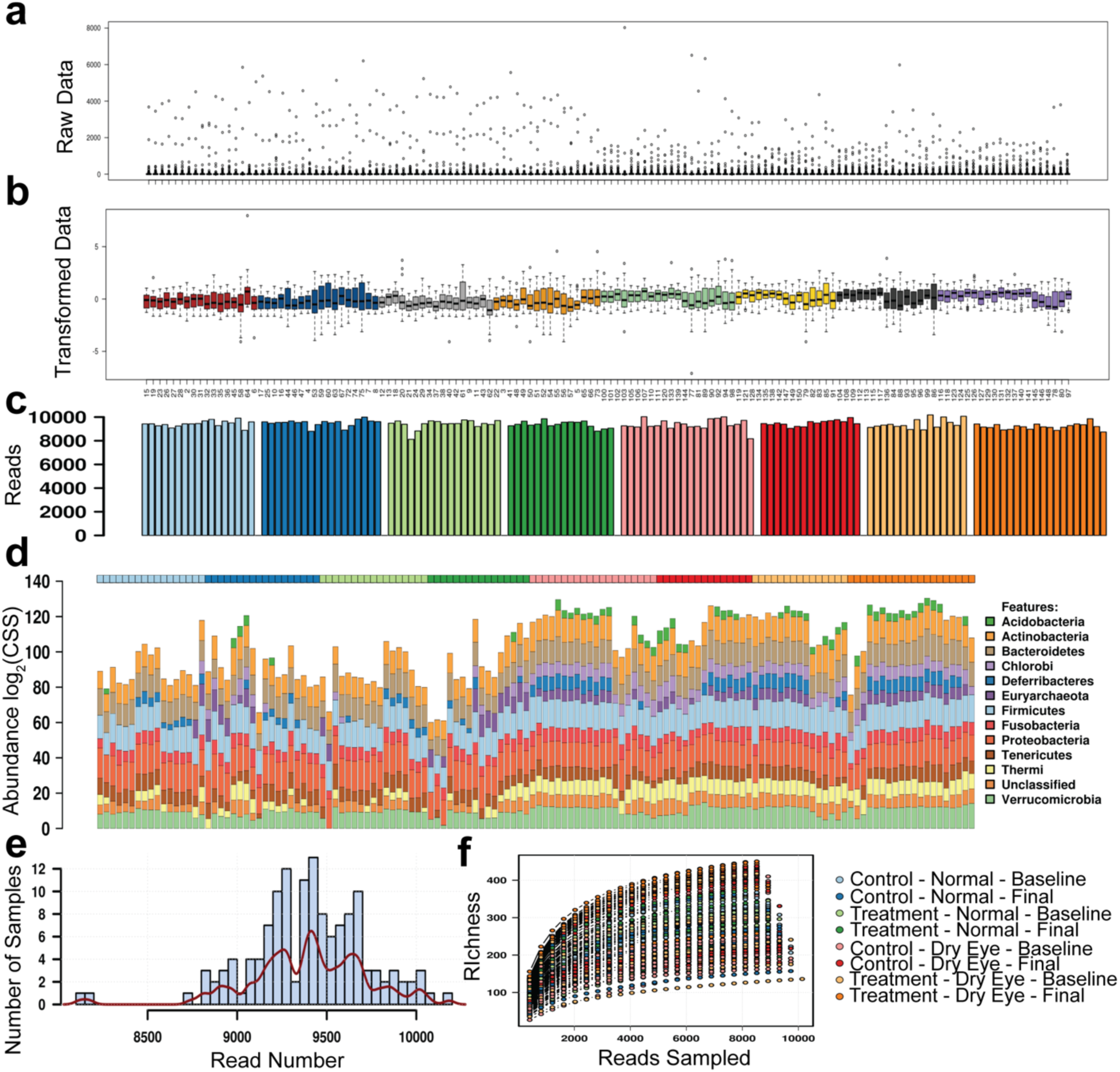
Read depth and quality are consistent across clinical groupings. (a) Raw sequencing data. (b) Log_2_-transformed relative abundance. (c) Sequencing depth. (d) Relative abundance at the phylum level. (e) Sequence reads per sample. (f) Read depth.

**Fig. S3.**
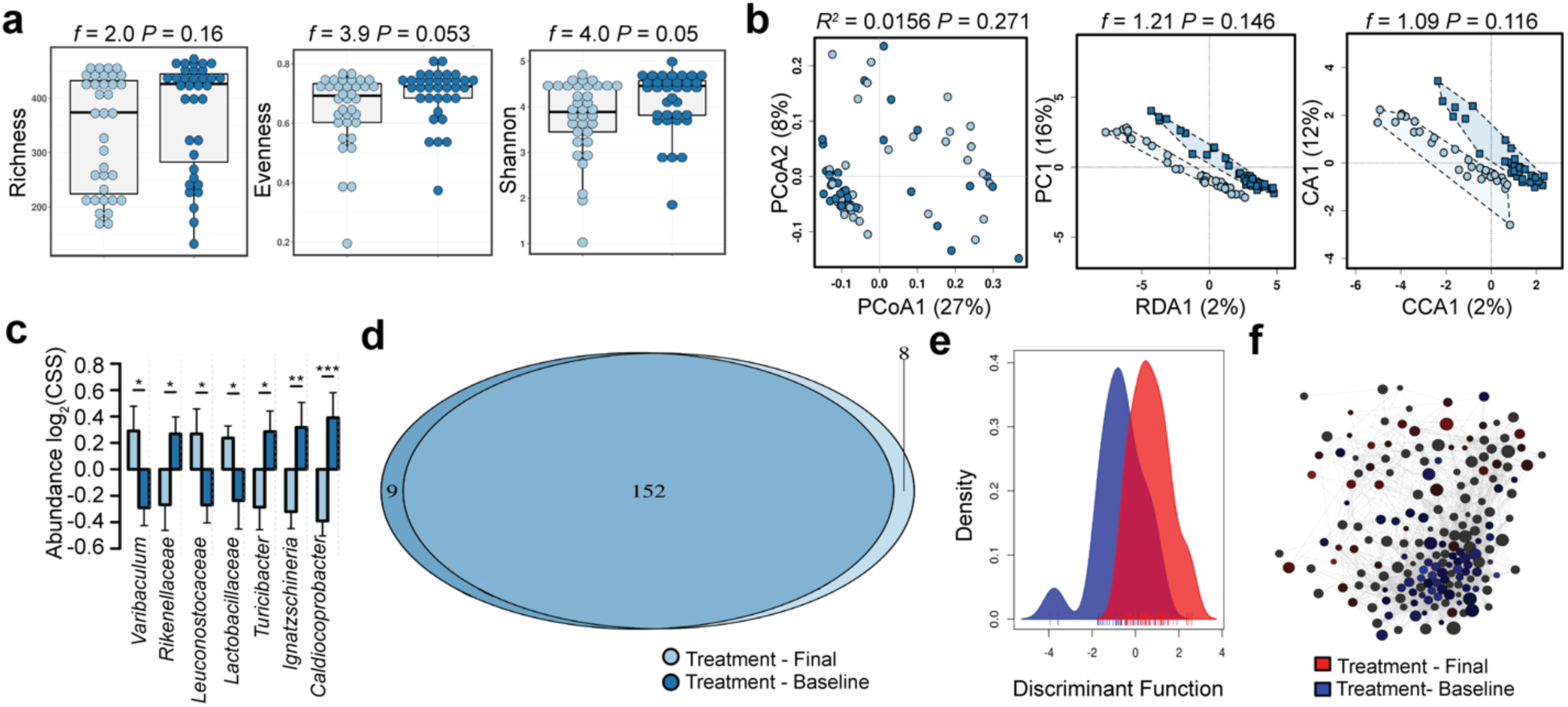
Limited changes occur in the dry eye microbiome with daily eye rinse. (a) Alpha diversity. (b) Beta diversity. (c) Relative abundance of bacterial genera. Two-way ANOVA, * *P* =0.05, ** *P* =0.01, *** *P* = 0.001. (d) Core microbiome analysis. (e) Discriminant analysis of principal components. (f) Spearman network analysis at the genus level.

**Fig. S4.**
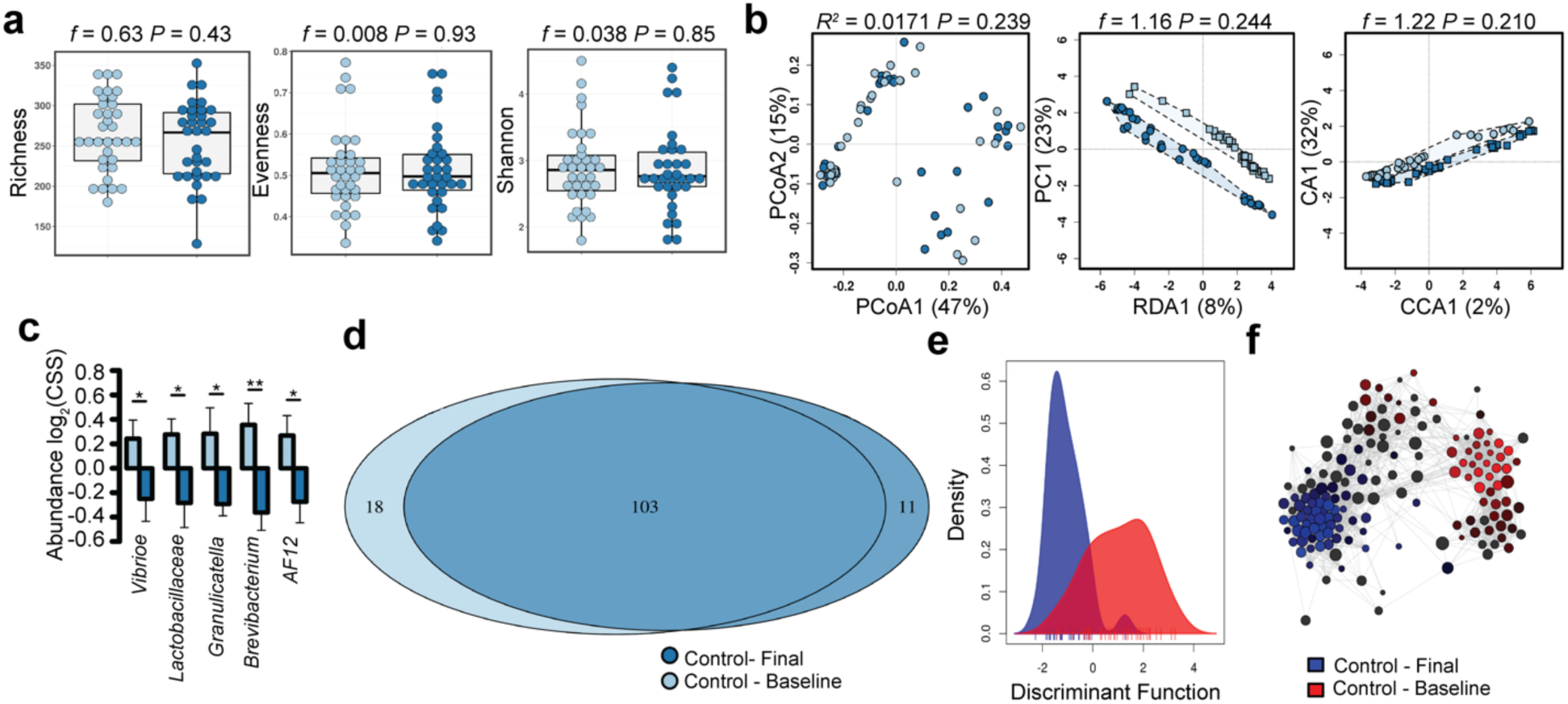
Limited changes occur in the normal eye microbiome with eye rinse. (a) Alpha diversity. (b) Beta diversity. (c) Relative abundance of bacterial orders. Two-way ANOVA, * *P* =0.05, ** *P* =0.01, *** *P* = 0.001. (d) Core microbiome analysis. (e) Discriminant analysis of principal components. (f) Spearman network analysis at the genus level.

**Fig. S5.**
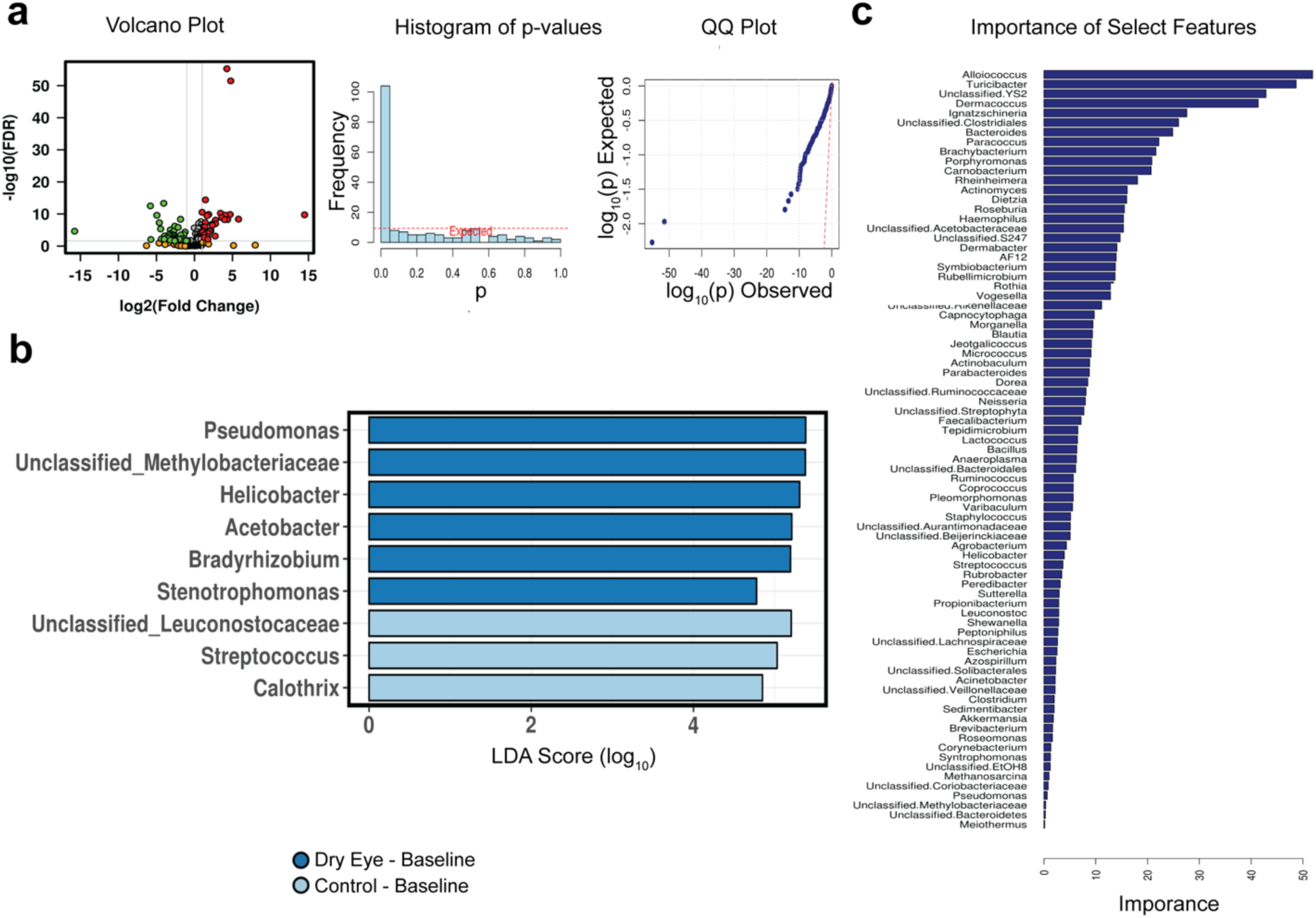
The acquisition of unique taxa separates the dry from the normal eye at baseline. (a) Negative binomial distribution (DESeq2 function). (b) Linear discriminant analysis of effect size (LEfSe). (c) Variable importance analysis of random forest classifier.

**Fig. S6.**
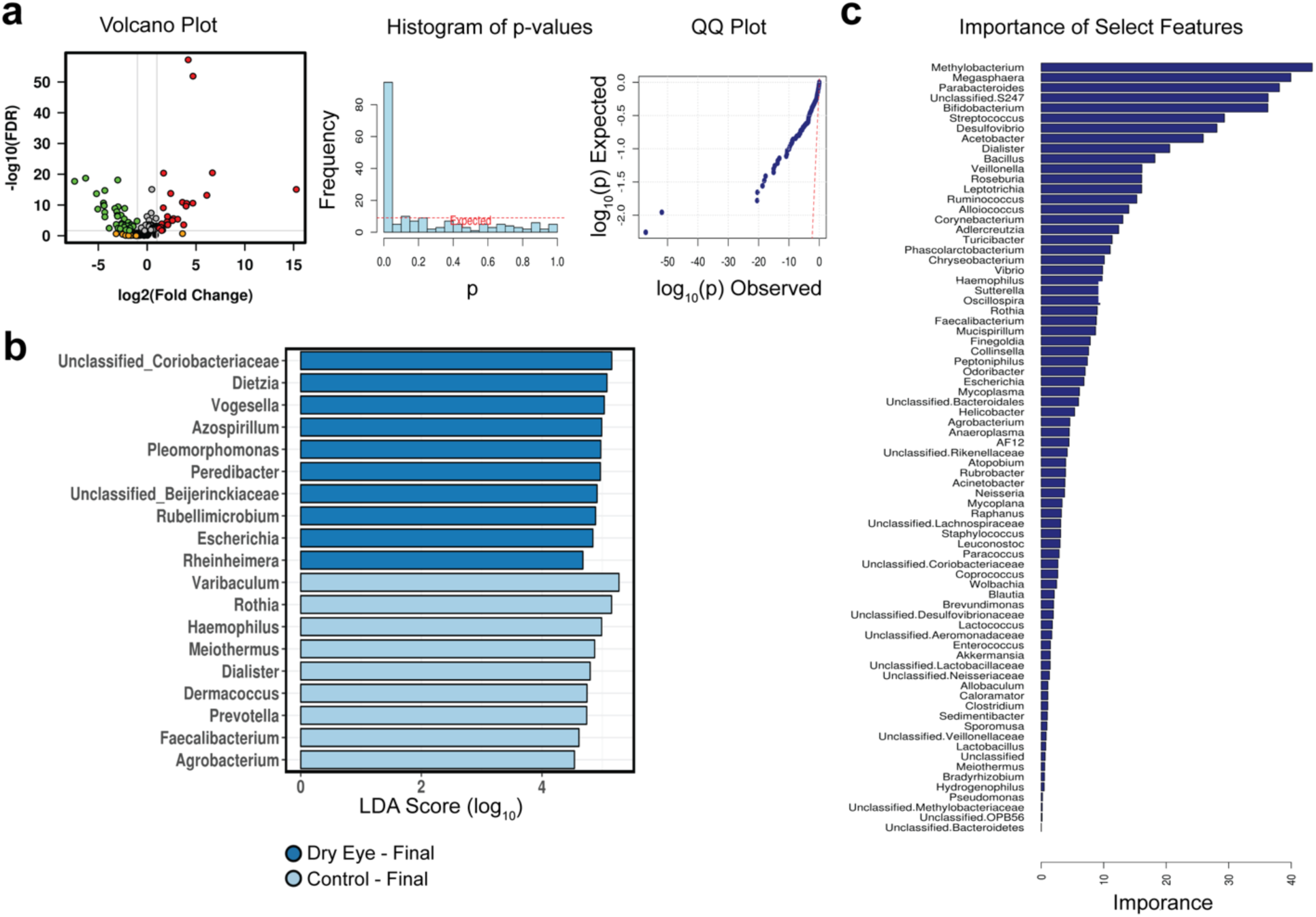
Additional unique taxa separate the dry from the normal eye after a month. (a) Negative binomial distribution (DESeq2 function). (b) Linear discriminant analysis of effect size (LEfSe). (c) Variable importance analysis of random forest classifier.

**Supplemental Table 1.**

|  | Normal |  | Dry Eye |  |
| --- | --- | --- | --- | --- |
|  | Normal | Mild | Moderate | Severe |
| **DEQ5** | ≤ 5 | ≤ 5 | ≥ 6 | ≥ 12 |
|  | All of the below: |  | At least one of the following: |  |
| **Phenol Red Thread, mm** | > 10, bilateral | > 10, bilateral | < 10, one eye | < 10, one eye |
| **NIKBUT, s** | > 5, bilateral | > 5, bilateral | < 10, one eye | < 5, one eye |
| **InflammaDry** | Negative, both eyes | Positive, one or both eyes | Positive, one or both eyes | Positive, one or both eyes |

**Supplemental Table 2.**
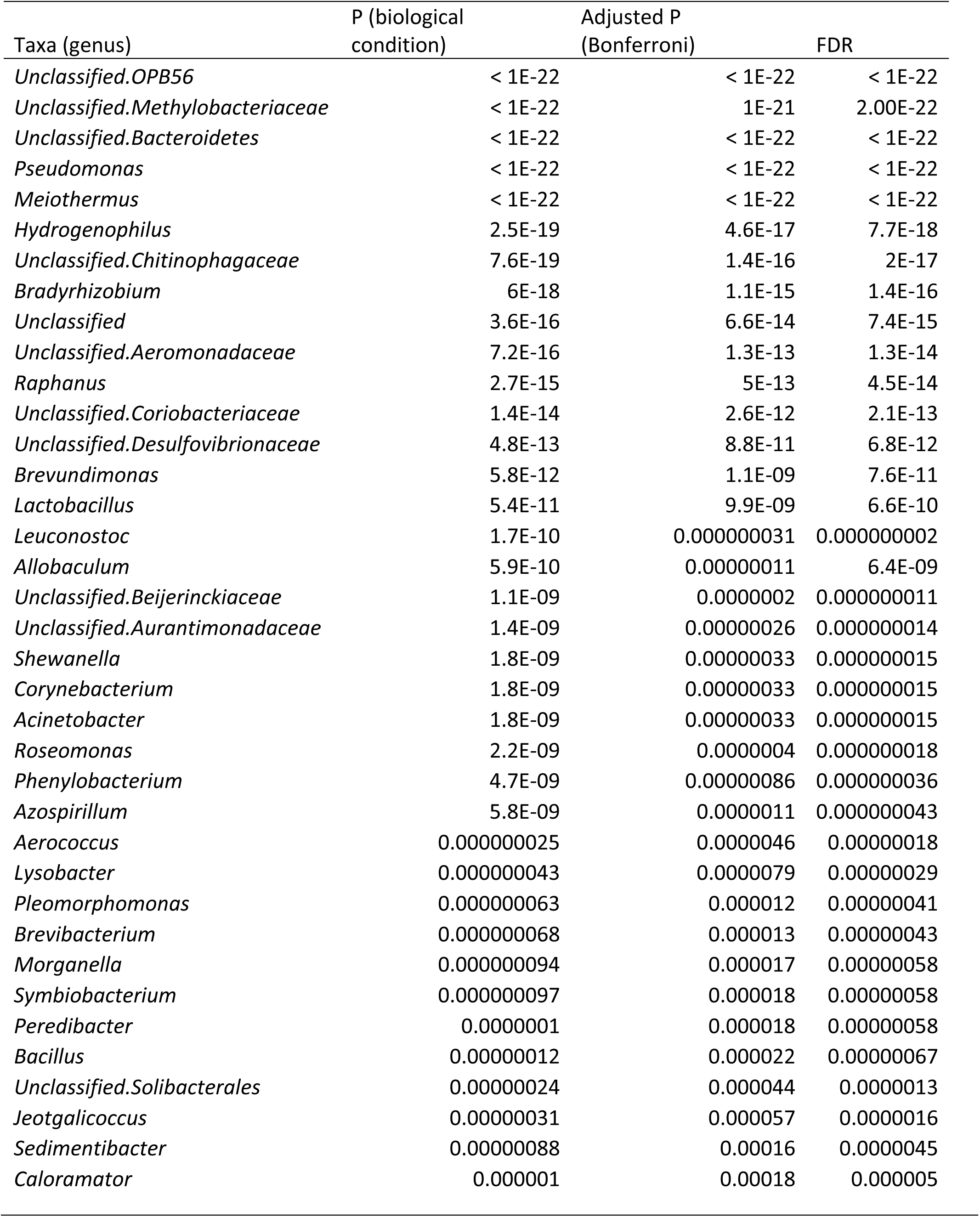
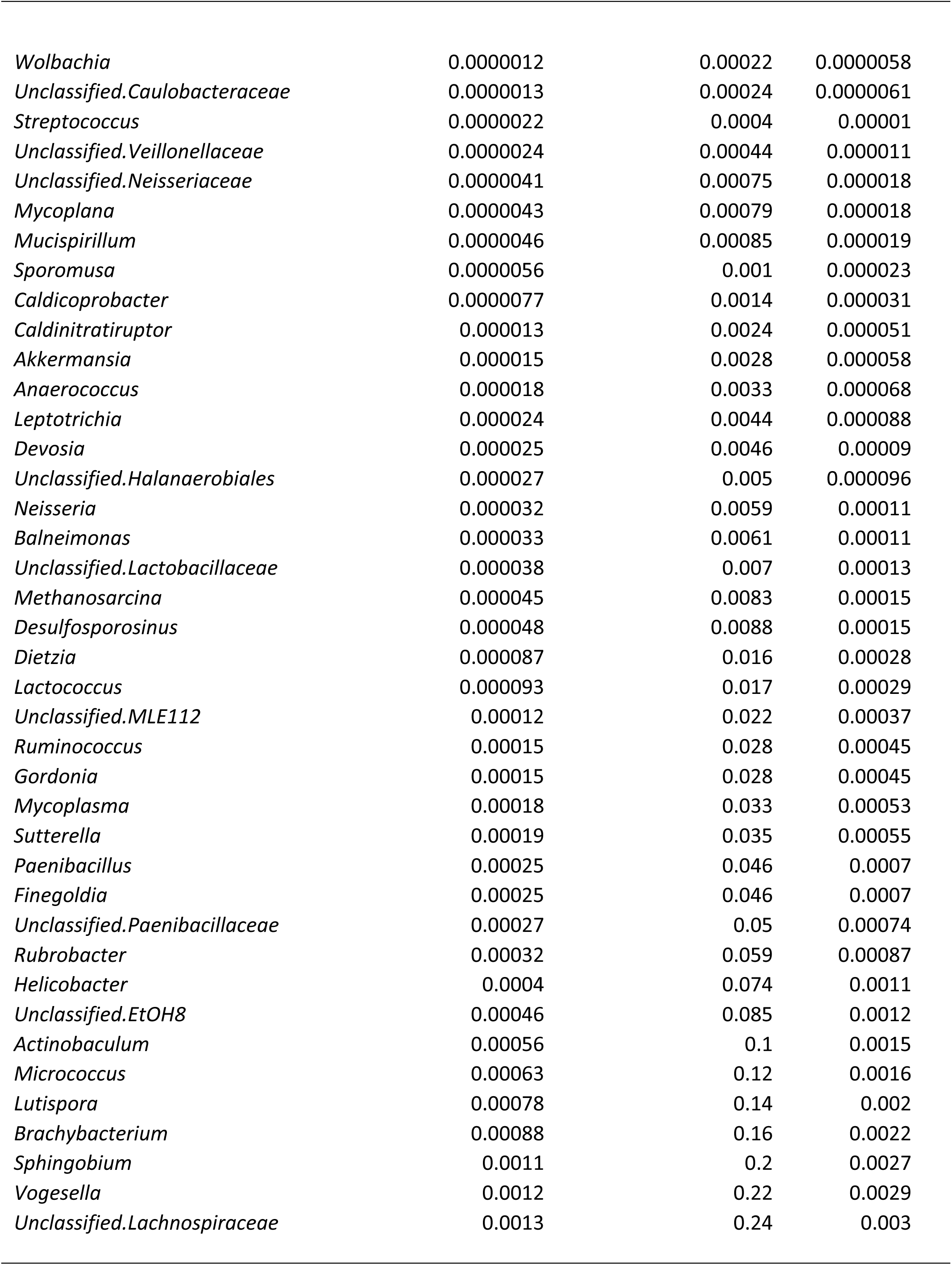
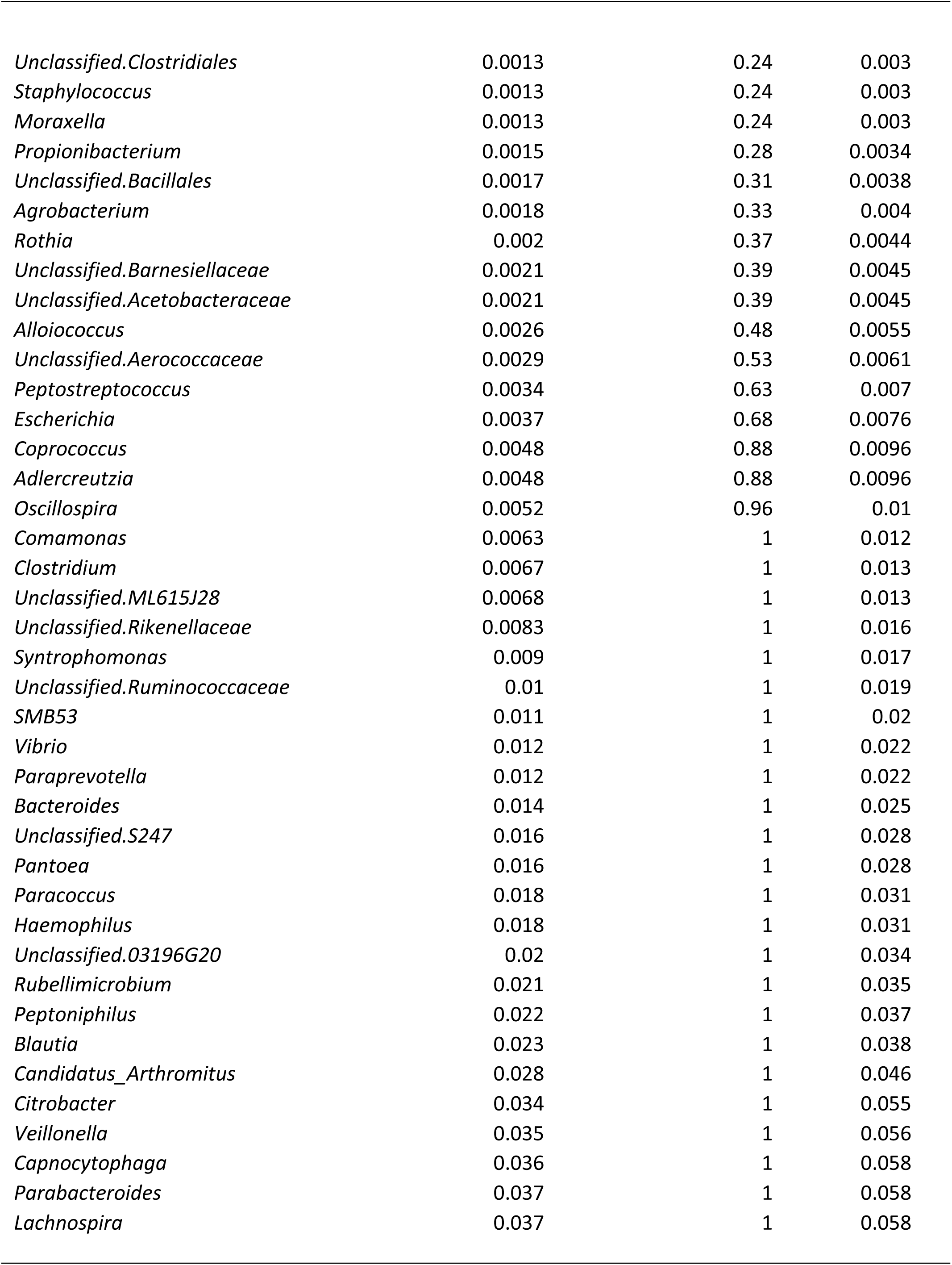
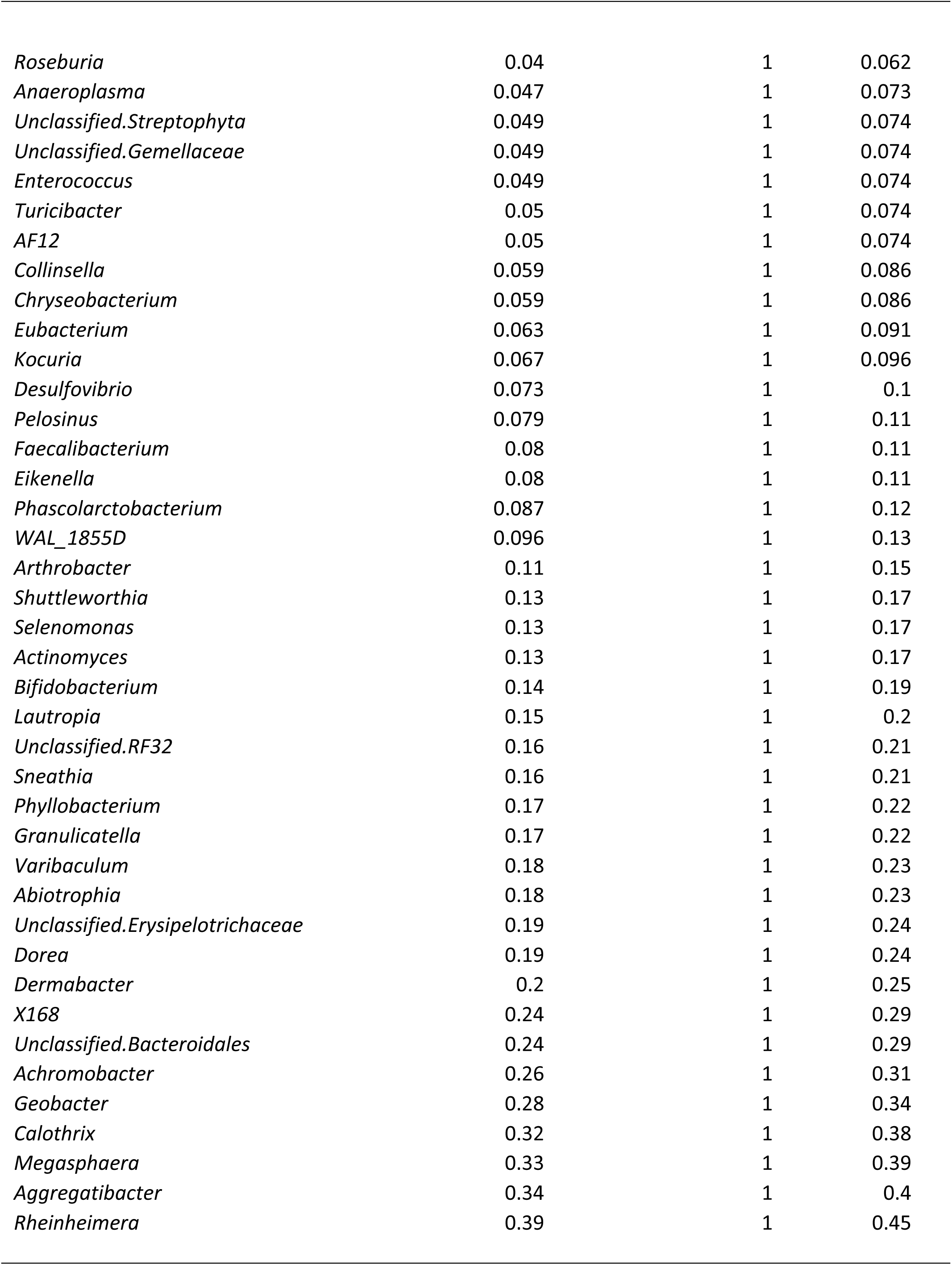
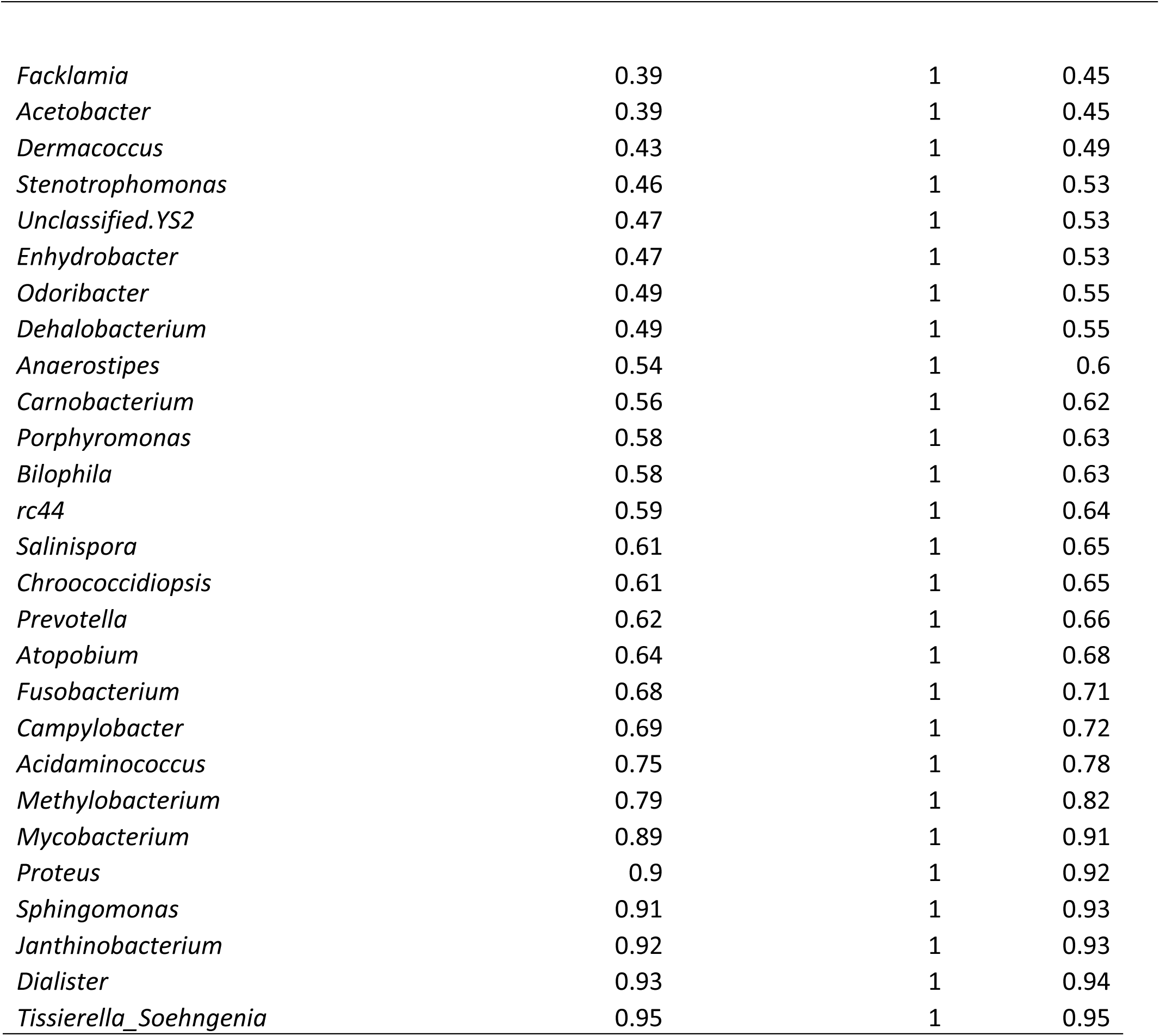

**Supplemental Table 3.**
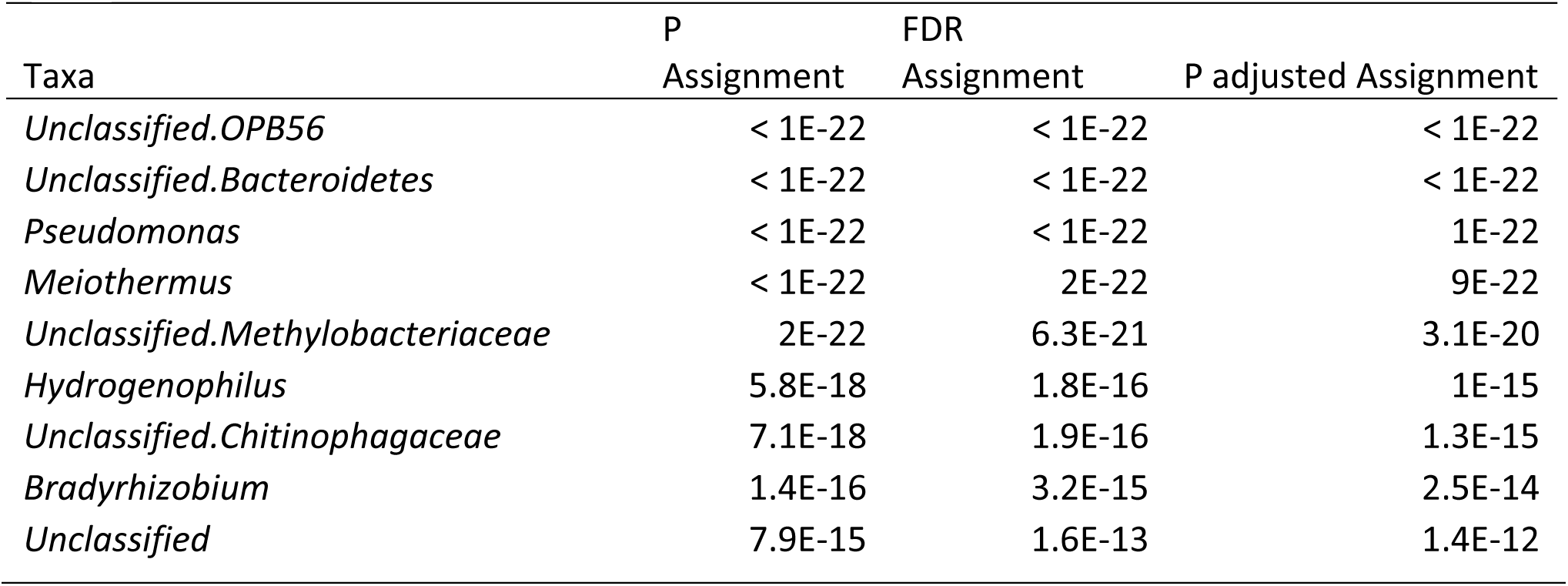
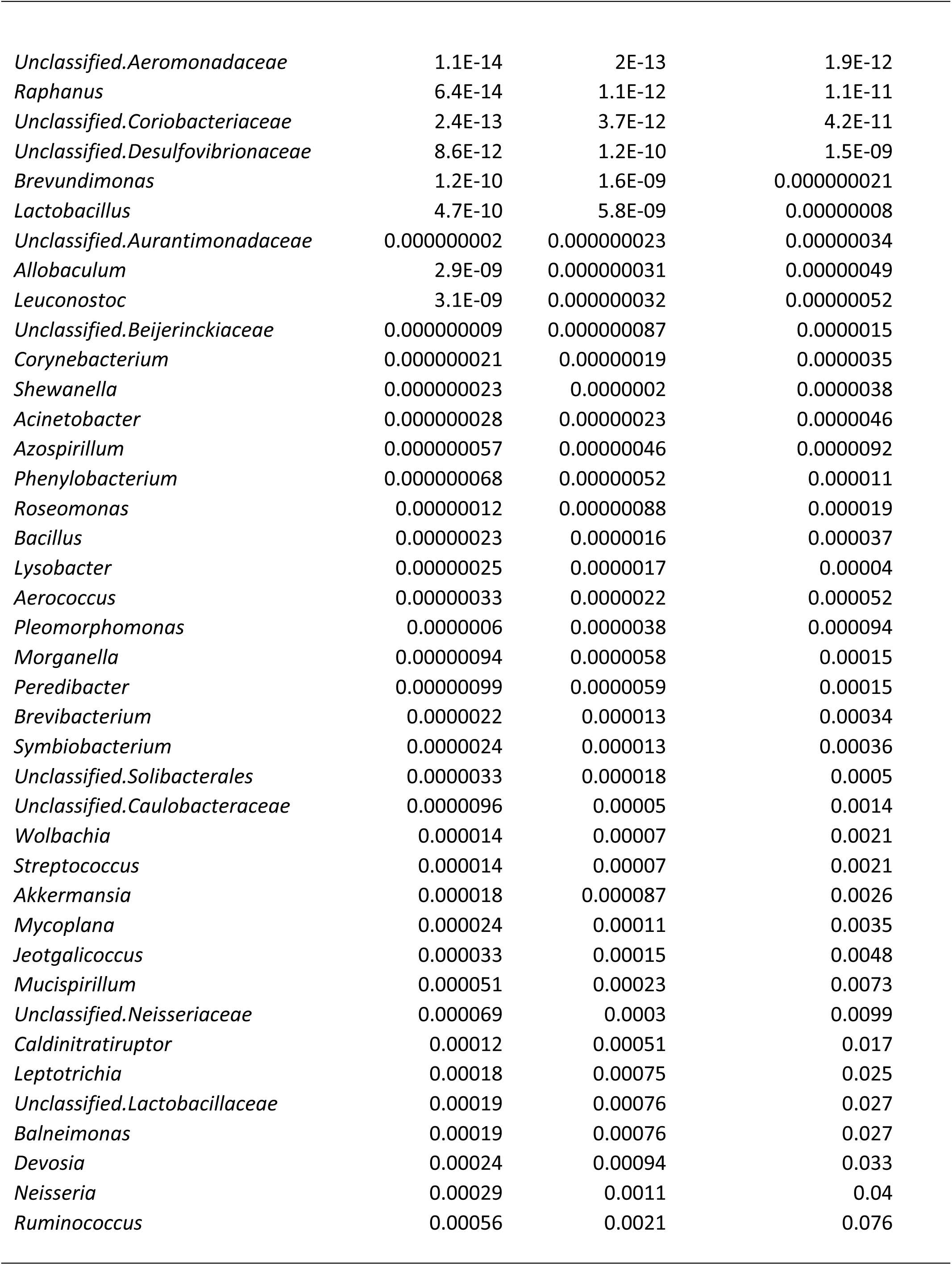
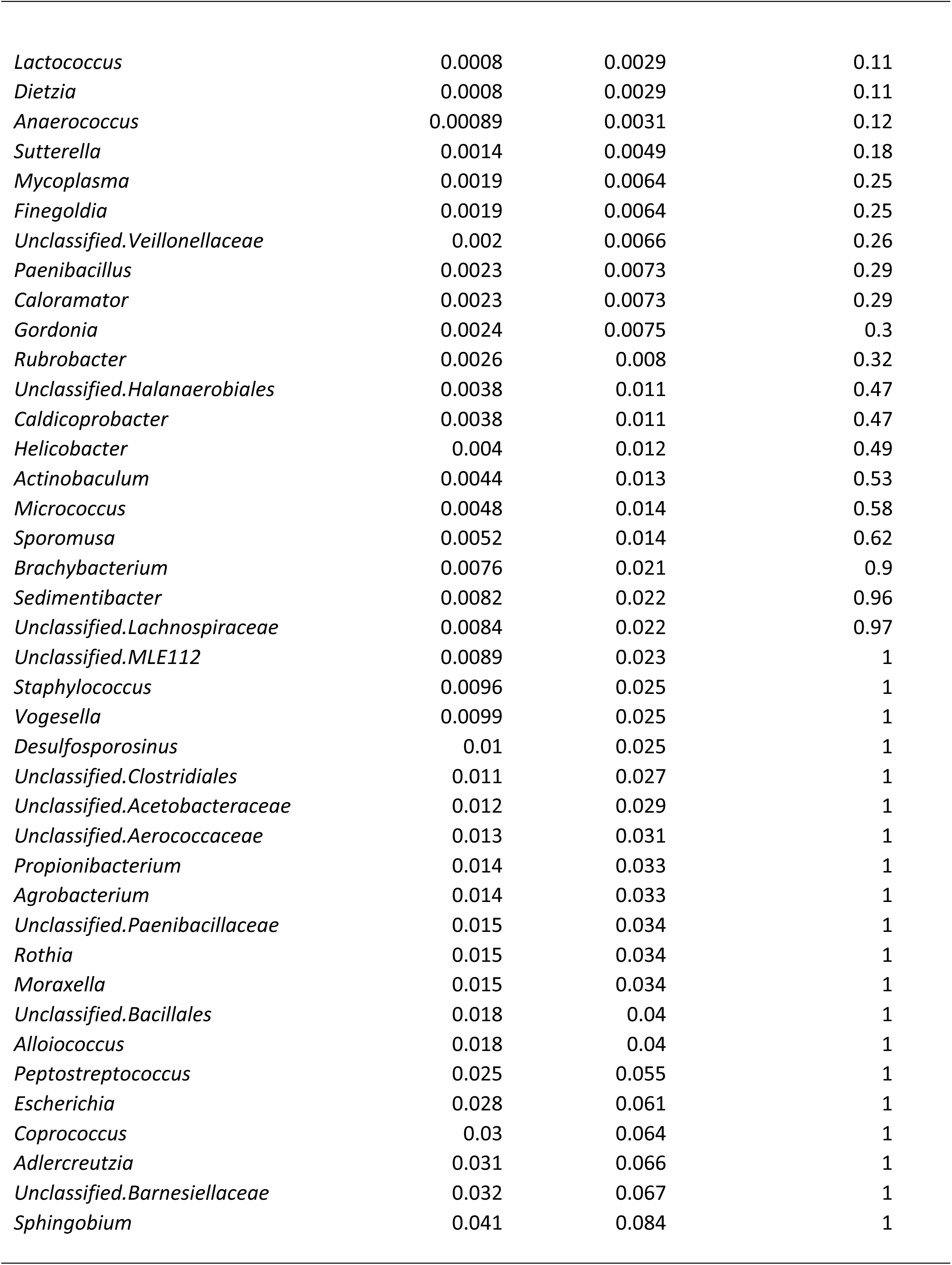
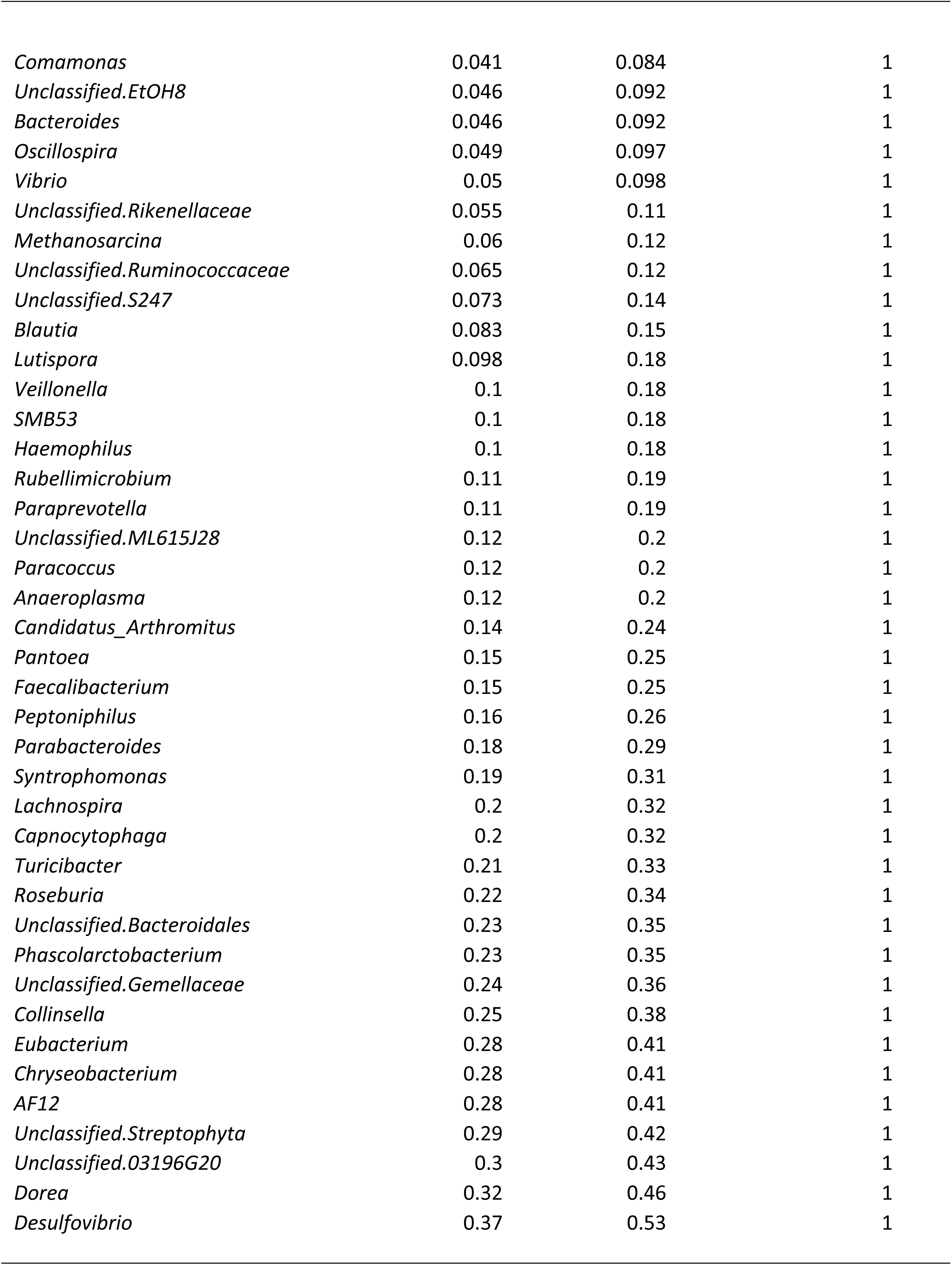
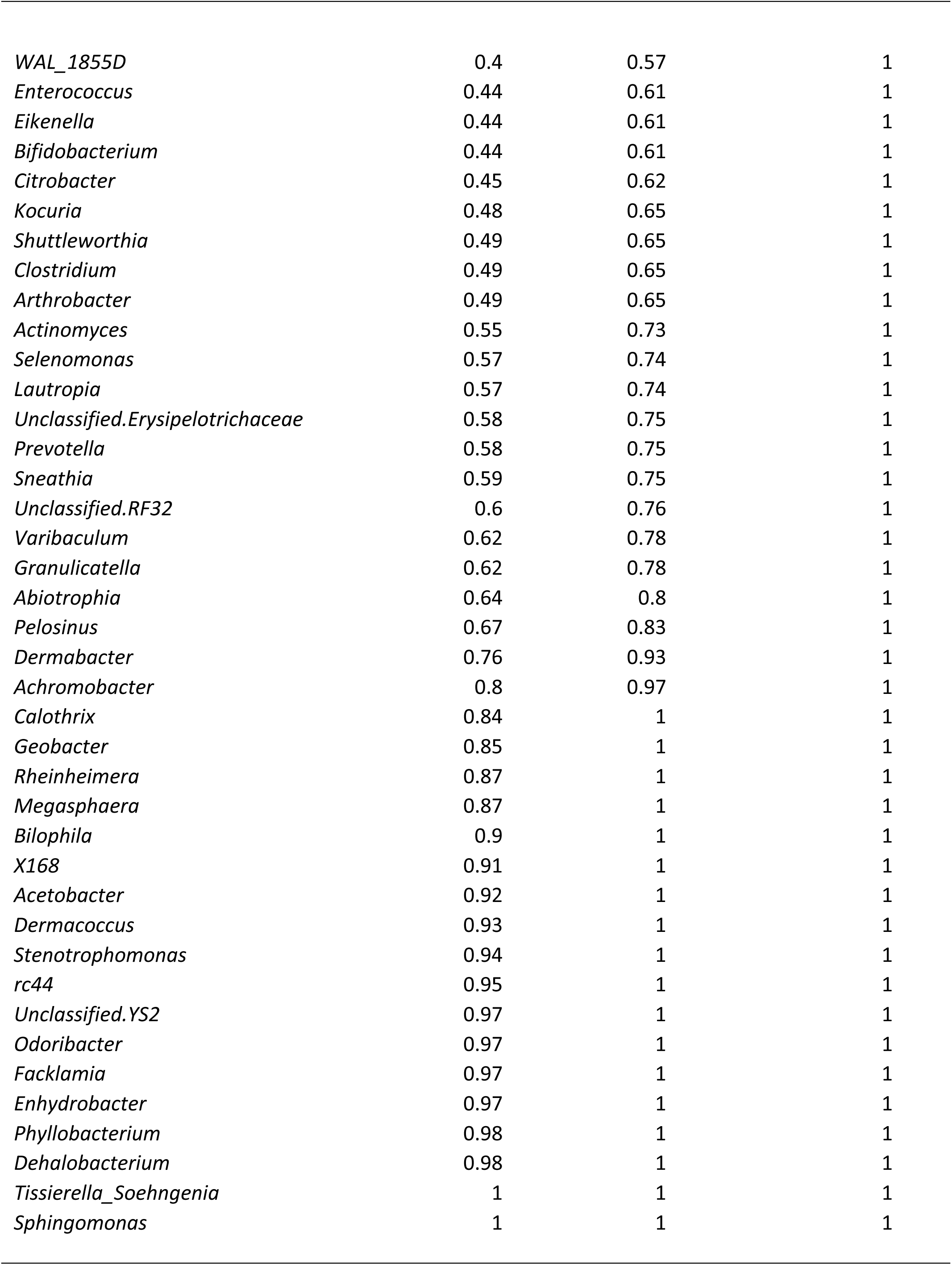
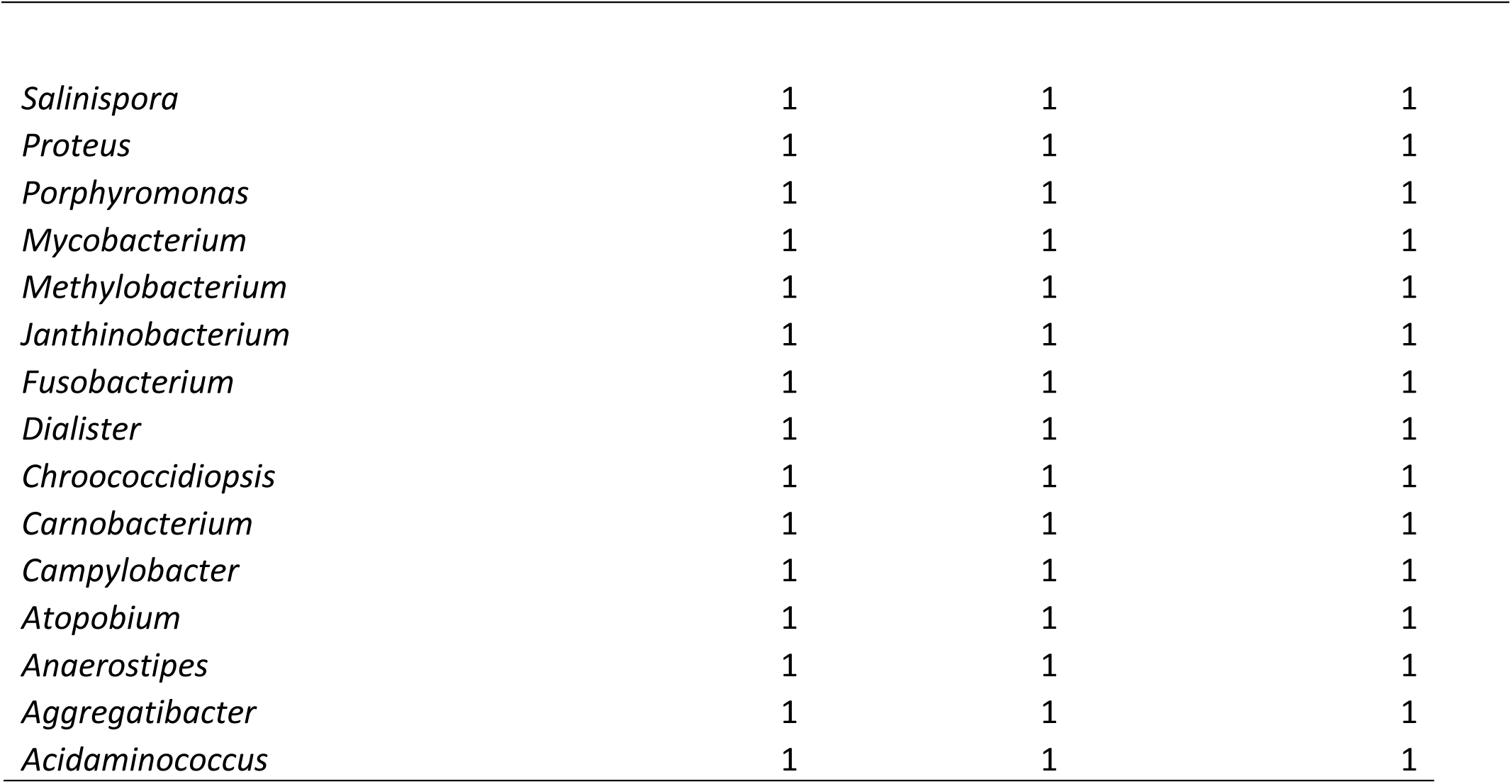

**Supplemental Table 4.**
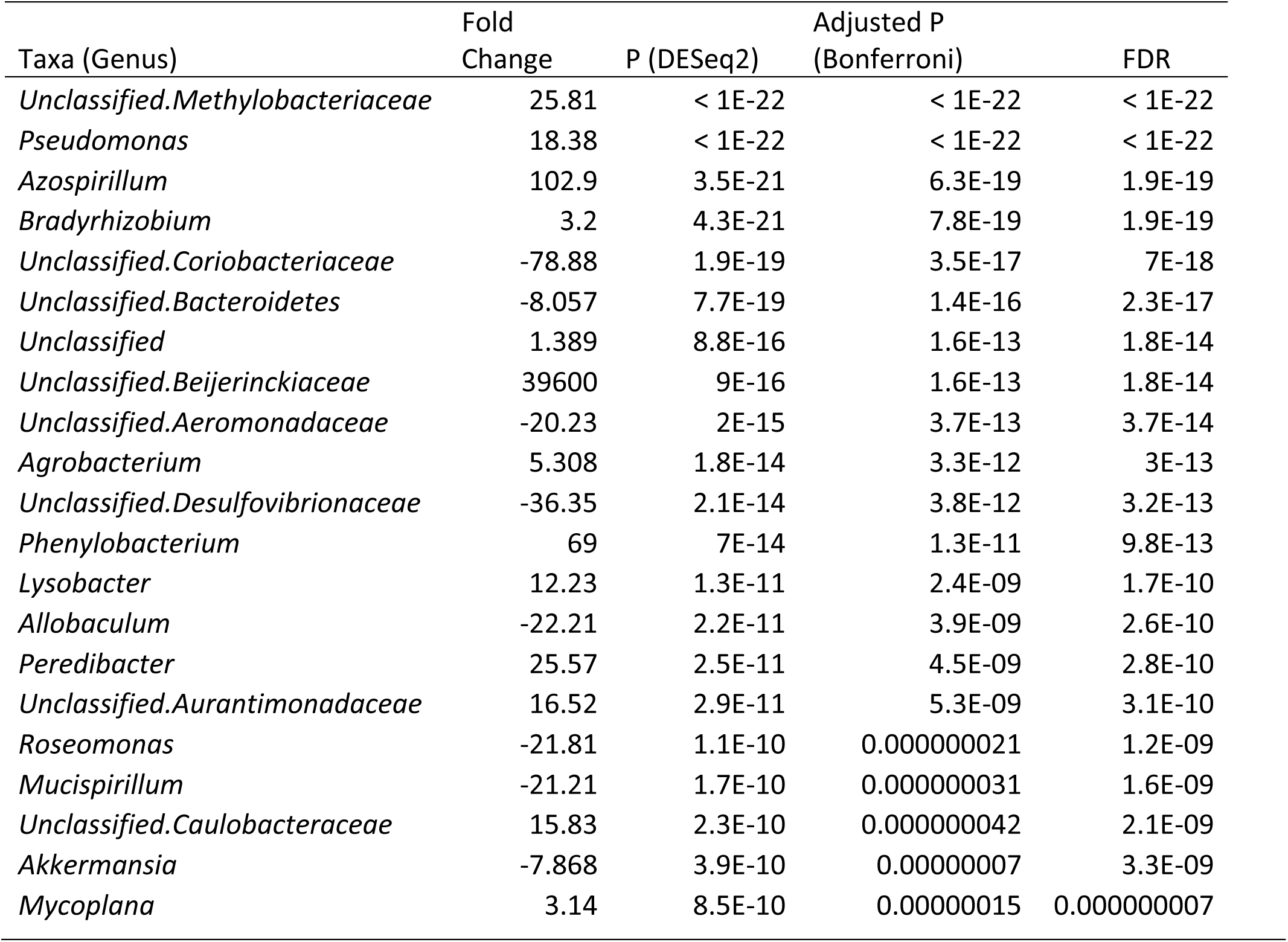
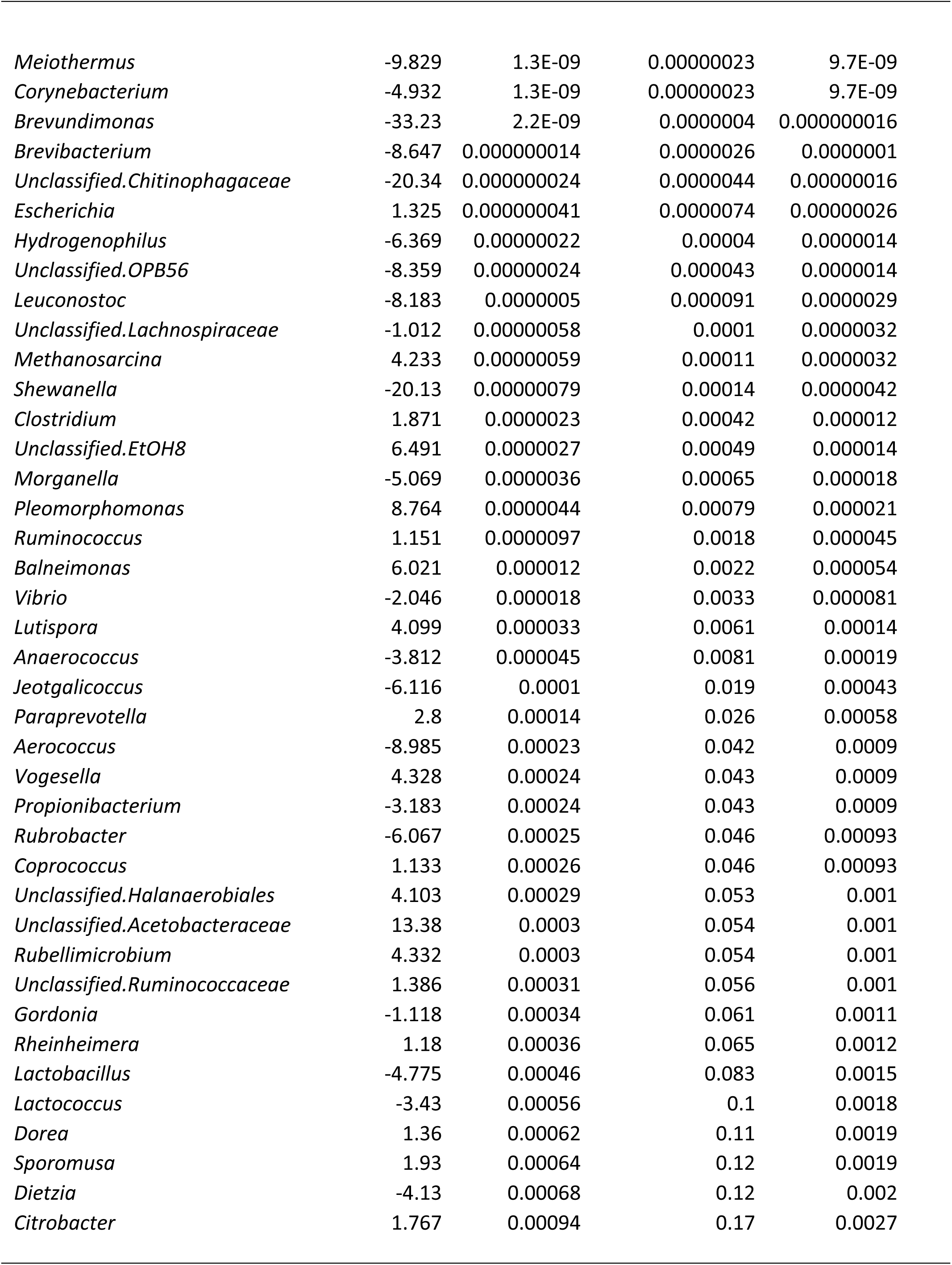
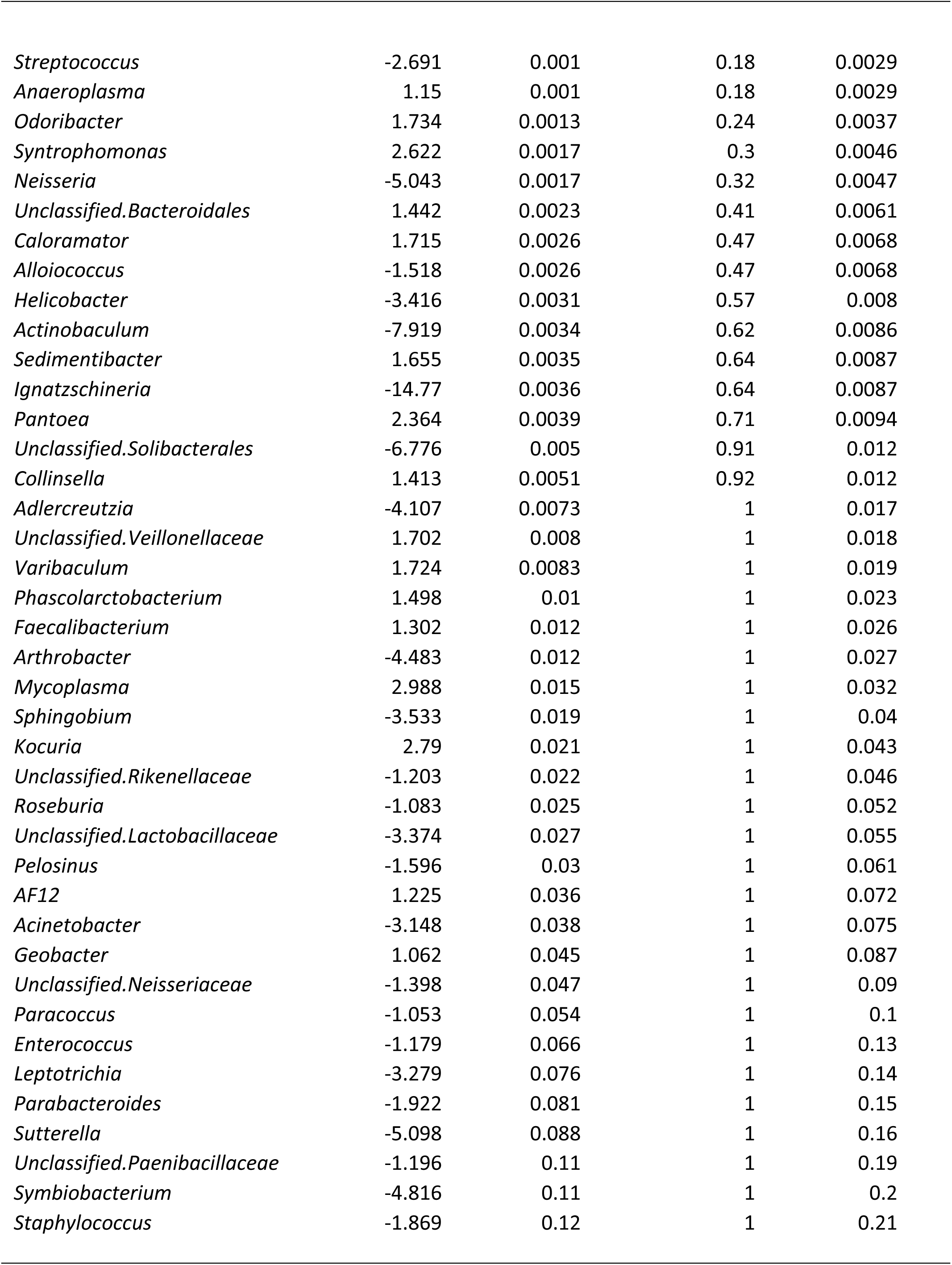
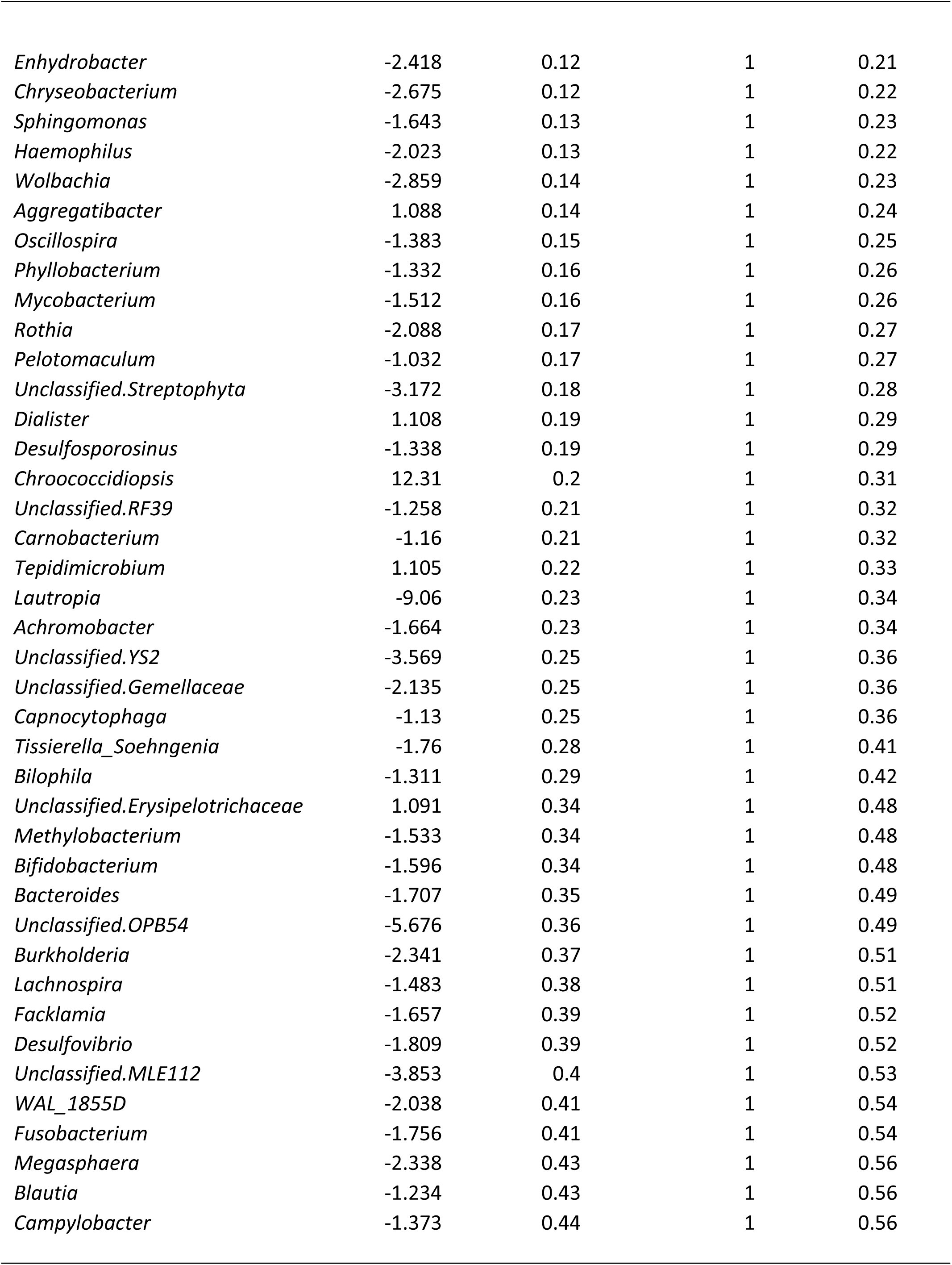
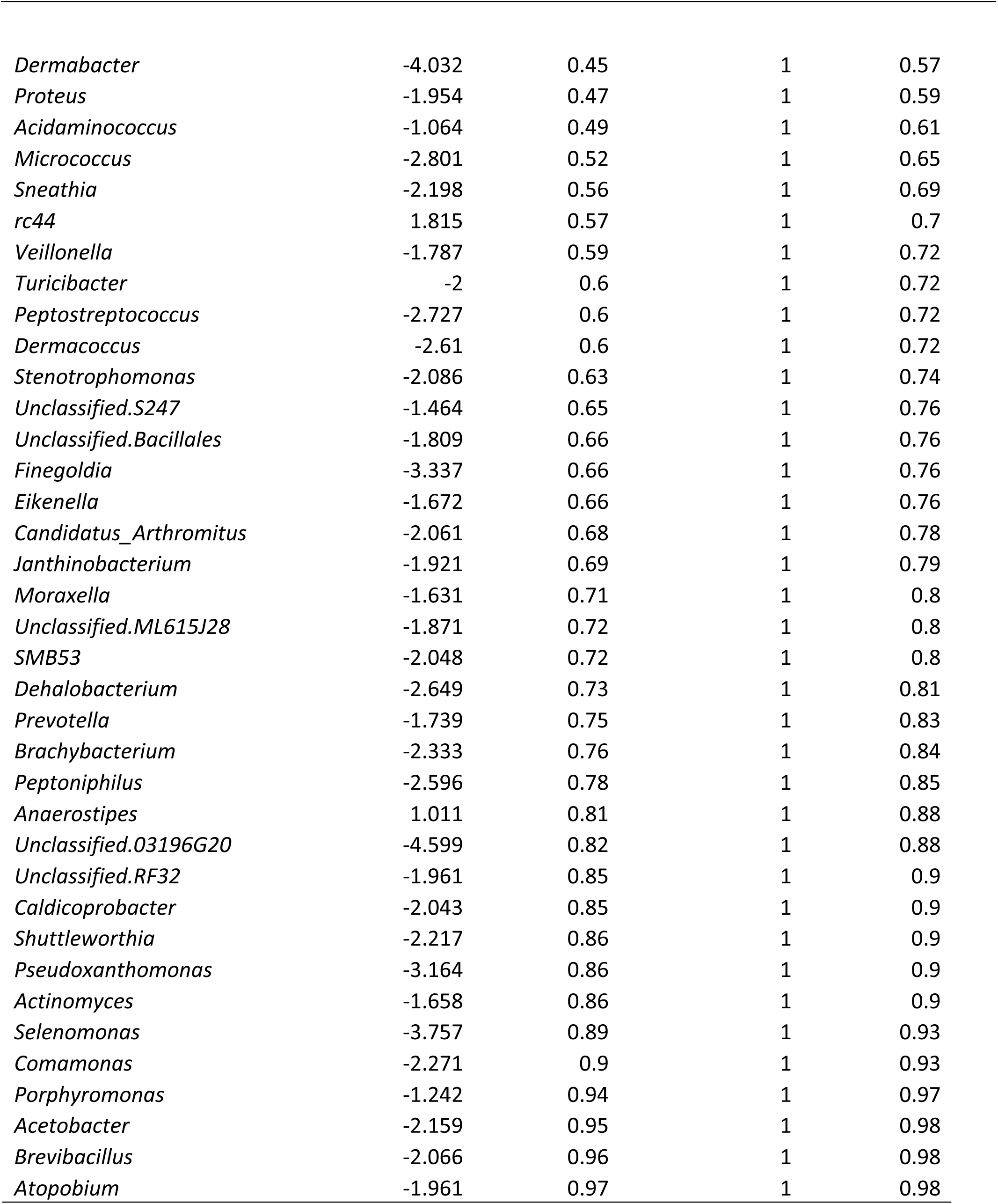

## References

1. Farrand, K. F., Fridman, M., Stillman, I. Ö. & Schaumberg, D. A. Prevalence of Diagnosed Dry Eye Disease in the United States Among Adults Aged 18 Years and Older. Am. J. Ophthalmol. 182, 90–98 (2017).

2. Stapleton, F. et al. TFOS DEWS II Epidemiology Report. The Ocular Surface 15, 334–365 (2017).

3. Chen, Y., Chauhan, S. K., Lee, H. S., Saban, D. R. & Dana, R. Chronic dry eye disease is principally mediated by effector memory Th17 cells. Mucosal Immunol 7, 38–45 (2014).

4. Heidari, M., Noorizadeh, F., Wu, K., Inomata, T. & Mashaghi, A. Dry Eye Disease: Emerging Approaches to Disease Analysis and Therapy. J Clin Med 8, 1439 (2019).

5. Gayton, J. L. Etiology, prevalence, and treatment of dry eye disease. Clin Ophthalmol 3, 405–412 (2009).

6. Cheng, H., Guan, X., Chen, D. & Ma, W. The Th17/Treg Cell Balance: A Gut Microbiota-Modulated Story. Microorganisms 7, 583 (2019).

7. Lee, Y. K., Menezes, J. S., Umesaki, Y. & Mazmanian, S. K. Proinflammatory T-cell responses to gut microbiota promote experimental autoimmune encephalomyelitis. Proc Natl Acad Sci U S A 108 Suppl 1, 4615–4622 (2011).

8. Willcox, M. D. P. Characterization of the normal microbiota of the ocular surface. Experimental Eye Research 117, 99–105 (2013).

9. Ozkan, J. et al. Temporal Stability and Composition of the Ocular Surface Microbiome. Sci Rep 7, 9880–11 (2017).

10. de Paiva, C. S. et al. Altered Mucosal Microbiome Diversity and Disease Severity in Sjögren Syndrome. Sci Rep 6, 23561–11 (2016).

11. Cavuoto, K. M., Banerjee, S. & Galor, A. Relationship between the microbiome and ocular health. The Ocular Surface 17, 384–392 (2019).

12. Baim, A. D., Movahedan, A., Farooq, A. V. & Skondra, D. The microbiome and ophthalmic disease. Exp. Biol. Med. (Maywood) 244, 419–429 (2019).

13. Narayanan, S., Redfern, R. L., Miller, W. L., Nichols, K. K. & McDermott, A. M. Dry Eye Disease and Microbial Keratitis: Is There a Connection? The Ocular Surface 11, 75–92 (2013).

14. Kelly, B. J. et al. Power and sample-size estimation for microbiome studies using pairwise distances and PERMANOVA. Bioinformatics 31, 2461–2468 (2015).

15. DeSantis, T. Z. et al. Greengenes, a chimera-checked 16S rRNA gene database and workbench compatible with ARB. Appl. Environ. Microbiol. 72, 5069–5072 (2006).

16. Glassman, S. I. & Martiny, J. B. H. Broadscale Ecological Patterns Are Robust to Use of Exact Sequence Variants versus Operational Taxonomic Units. mSphere 3, 5409 (2018).

17. Paulson, J. N., Stine, O. C., Bravo, H. C. & Pop, M. Robust methods for differential abundance analysis in marker gene surveys. Nat. Methods 10, 1200–1202 (2013).

18. Postnikoff, C. K., Huisingh, C., McGwin, G. & Nichols, K. K. Leukocyte Distribution in the Open Eye Tears of Normal and Dry Eye Subjects. Curr. Eye Res. 43, 1253–1259 (2018).

19. Ahern, P. P. & Maloy, K. J. Understanding immune-microbiota interactions in the intestine. Immunology 22, 283 (2019).

20. Postnikoff, C. K. & Nichols, K. K. Neutrophil and T-Cell Homeostasis in the Closed Eye. Invest. Ophthalmol. Vis. Sci. 58, 6212–6220 (2017).

21. Shin, H. et al. Changes in the Eye Microbiota Associated with Contact Lens Wearing. mBio 7, e00198 (2016).

22. Doan, T. et al. Paucibacterial Microbiome and Resident DNA Virome of the Healthy Conjunctiva. Invest. Ophthalmol. Vis. Sci. 57, 5116–5126 (2016).

23. Graham, J. E. et al. Ocular pathogen or commensal: a PCR-based study of surface bacterial flora in normal and dry eyes. Invest. Ophthalmol. Vis. Sci. 48, 5616–5623 (2007).

24. Albietz, J. M. & Lenton, L. M. Effect of antibacterial honey on the ocular flora in tear deficiency and meibomian gland disease. Cornea 25, 1012–1019 (2006).

25. Costello, E. K., Stagaman, K., Dethlefsen, Les, Bohannan, B. J. M. & Relman, D. A. The Application of Ecological Theory Toward an Understanding of the Human Microbiome. Science 336, 1255–1262 (2012).

26. McDermott, A. M. Antimicrobial compounds in tears. Experimental Eye Research 117, 53–61 (2013).

27. Yin, V. T. et al. Antibiotic Resistance of Ocular Surface Flora With Repeated Use of a Topical Antibiotic After Intravitreal Injection. JAMA Ophthalmol 131, 456–461 (2013).

28. Iovieno, A. et al. Preliminary evidence of the efficacy of probiotic eye-drop treatment in patients with vernal keratoconjunctivitis. Graefes Arch. Clin. Exp. Ophthalmol. 246, 435–441 (2008).

29. Round, J. L. & Palm, N. W. Causal effects of the microbiota on immune-mediated diseases. Sci Immunol 3, eaao1603 (2018).

30. Willis, K. A. et al. Perinatal maternal antibiotic exposure augments lung injury in offspring in experimental bronchopulmonary dysplasia. American Journal of Physiology - Lung Cellular and Molecular Physiology 3, 21 (2019).

31. Dolma, K. et al. Effects of Hyperoxia on Alveolar and Pulmonary Vascular Development in Germ Free Mice. American Journal of Physiology - Lung Cellular and Molecular Physiology 295, L86 (2019).

32. Wang, Y., Chen, H., Xia, T. & Huang, Y. Characterization of fungal microbiota on normal ocular surface of humans. Clin. Microbiol. Infect. (2019). doi:10.1016/j.cmi.2019.05.011

33. Willis, K. A. et al. Fungi form interkingdom microbial communities in the primordial human gut that develop with gestational age. FASEB J. 33, 12825–12837 (2019).

34. Lal, C. V. et al. The airway microbiome at birth. nature.com

35. Chalmers, R. L., Begley, C. G. & Caffery, B. Validation of the 5-Item Dry Eye Questionnaire (DEQ-5): Discrimination across self-assessed severity and aqueous tear deficient dry eye diagnoses. Contact Lens and Anterior Eye 33, 55–60 (2010).

36. Schiffman, R. M., Christianson, M. D., Jacobsen, G., Hirsch, J. D. & Reis, B. L. Reliability and Validity of the Ocular Surface Disease Index. Arch Ophthalmol 118, 615–621 (2000).

37. Miller, W. L. et al. A Comparison of Tear Volume (by Tear Meniscus Height and Phenol Red Thread Test) and Tear Fluid Osmolality Measures in Non-Lens Wearers and in Contact Lens Wearers. Eye & Contact Lens 30, 132–137 (2004).

38. Harris, P. A. et al. The REDCap consortium: Building an international community of software platform partners. J Biomed Inform 95, 103208 (2019).

39. Postnikoff, C. K., Huisingh, C., McGwin, G. & Nichols, K. K. Leukocyte Distribution in the Open Eye Tears of Normal and Dry Eye Subjects. Curr. Eye Res.43, 1253–1259 (2018).

40. Postnikoff, C. K., ophthalmology, K. N. I.2017. Neutrophil and T-cell homeostasis in the closed eye. iovs.arvojournals.org

41. Gorbet, M., Postnikoff, C. & Williams, S. The Noninflammatory Phenotype of Neutrophils From the Closed-Eye Environment: A Flow Cytometry Analysis of Receptor Expression. Invest. Ophthalmol. Vis. Sci. 56, 4582–4591 (2015).

42. Caporaso, J. G. et al. QIIME allows analysis of high-throughput community sequencing data. Nat. Methods 7, 335–336 (2010).

43. Zakrzewski, M. et al. Calypso: a user-friendly web-server for mining and visualizing microbiome-environment interactions. Bioinformatics 33, 782–783 (2017).

44. Reese, A. T. & Dunn, R. R. Drivers of Microbiome Biodiversity: A Review of General Rules, Feces, and Ignorance. mBio 9, 734 (2018).

45. Paliy, O. & Shankar, V. Application of multivariate statistical techniques in microbial ecology. Mol. Ecol. 25, 1032–1057 (2016).

46. Anderson, M. J. Permutational Multivariate Analysis of Variance (PERMANOVA). 18, 1–15 (American Cancer Society, 2014).

